# Genome-wide association meta-analyses over one million individuals identify 54 loci associated with urinary incontinence and its subtypes

**DOI:** 10.64898/2026.03.12.26346952

**Authors:** Elisa Moreno, Nikhil Arora, Emily Bertucci-Richter, Reyhane Eghtedarian, Aristomo Andries, Laurent F. Thomas, Julie Horn, Signe N. Stafne, Sweta Pathak, FinnGen, Snehal Patil, Dagny Gulla, Hanna M. Ollila, David M. Evans, Giulia M. Ippolito, Wei Zhou, Eivind Coward, Kristian Hveem, Ida Surakka, Marta R. Moksnes, Brooke N. Wolford, Ben M. Brumpton

## Abstract

Urinary incontinence (UI) markedly reduces quality of life, and its genetic basis remains to be elucidated. Here, we performed GWAS meta-analyses of four UI phenotypes, using hospital-based data from 1,045,436 individuals from the Trøndelag Health Study (HUNT), UK Biobank, FinnGen, and the Michigan Genomics Initiative, and self-reported UI from 56,957 females from HUNT and the Nurses’ Health Study.

We identified 54 (52 novels) genetic loci associated with at least one UI phenotype. Analyses revealed tissue-specific involvement, with connective and muscular tissues for stress UI, and neural tissues for urge UI, and supported key roles of SUI-prioritized genes in the extracellular matrix organization and muscle function. Further, genetic variations contributes to UI through inflammation, immune, and aging pathways. We identified smoking, higher BMI, higher parity, and pelvic organ prolapse as causal risk factors for UI in females, and benign prostatic hyperplasia in males. Our findings identify key genetic factors, tissues, pathways, and causal underlying UI.

## Introduction

Urinary incontinence (UI) is the involuntary leakage of urine and is strongly associated with poor quality of life (1). Most population studies report UI prevalence ranging from 25-45% in adult women (considering occasional leakage) and 1%-39% in males, with prevalences increasing with age (2). However, its burden tends to be underestimated at the population level due to stigma and lack of recognition as a medical condition (3). Two main subtypes of UI are 1) stress urinary incontinence (SUI) defined as urine leakage upon coughing, sneezing, laughing or during physical exertion and 2) urge urinary incontinence (UUI) defined as a urine leakage associated with a sudden need to void that is difficult to defer (4,5). Both subtypes can co-exist, in which case the condition is referred to as mixed urinary incontinence (MUI). Symptoms can be ameliorated by treatment and diverse clinical trials are ongoing (3).

Urinary continence is regulated and maintained through a complex coordination of smooth and skeletal muscles, nerves, bladder mucosa, connective tissues, the pelvic floor, and both the central and peripheral nervous systems (3,6). SUI primarily occurs when bladder pressure exceeds urethral pressure, often from increased intra-abdominal pressure, insufficient urethral support or sphincter dysfunction (3). In contrast, UUI is mainly characterized by detrusor muscle overactivity, with proposed mechanisms including muscle hyperexcitability, denervation, poor detrusor compliance and bladder hypersensitivity (3). However, UI etiology is far from fully understood. Despite heritability estimates of 39-63% from twin studies for SUI and 42-51% for UUI (7), genome-wide association studies (GWASs) have only identified 12 loci at the genome-wide significance threshold (p-value < 5×10^-8^) (8–10) and replicated two (9). In addition, the six GWASs of UI are limited by small sample sizes (n=1,507 to 25,685), few results replicated, and only one GWAS included males (8–13). Additionally, these studies did not include downstream analyses such as phenome-wide association studies (PheWASs) to explore pleiotropy and comorbidities, and Mendelian randomization (MR) analyses to assess causal risk factors and guide prevention. Observational studies suggest smoking status (14) and body mass index (BMI) (15,16) as common risk factors for UI, in addition to parity (14,17) and pelvic organ prolapse (PoP) for females, and benign prostatic hyperplasia (BPH) for males (15). However, it is unclear the extent to which these observational associations represent causal effects of the risk factors or the influence of confounding variables.

Here, we performed GWAS meta-analyses of four UI phenotypes (any type of UI (AUI), SUI, UUI and MUI) first using International Classification of Diseases (ICD) codes in up to 1,045,494 individuals from the Trøndelag Health Study (HUNT), UK Biobank, FinnGen and the Michigan Genomic Initiative (MGI), and secondly using self-reported data from up to 56,957 females participants in HUNT and the Nurses’ Health Study (NHS). Due to potential sex differences in pathological mechanisms, we also conducted sex-specific analyses. We performed PheWAS using summary statistics from UK Biobank, and calculated genetic correlations between UI and other publicly available phenotypes. Furthermore, we created a polygenic risk score (PRS) and assessed the variance explained in HUNT. Finally, we performed MR to test the causal effect of six suggested risk factors on UI. Identifying novel UI loci, validating previous findings and conducting additional downstream analyses can unravel biological pathways underlying the conditions, suggest drug targets for clinical trials and potentially improve prevention.

## Results

### GWAS genetic discovery

We performed GWAS meta-analyses of ICD code-based definitions (UI-ICD: AUI-ICD, SUI-ICD, UUI-ICD, MUI-ICD) across four population-based cohorts linked with hospital-based data, encompassing up to 1,045,436 individuals (58,918 cases, 986,518 controls) (**Figure 1; Supplementary Table 1; Supplementary Table 2**) and GWAS meta-analyses of self-reported questionnaire (UI-srq: AUI-srq, SUI-srq, UUI-srq, MUI-srq) including up to 56,957 females (23,392 cases, 33,565 controls) (**Supplementary Table 1; Supplementary Table 3**). The inflation factor (*λ*) at MAF>0.01 ranged between 0.987 and 1.13 (**Supplementary Table 4**), and the linkage disequilibrium score (LDSC) regression intercepts were below 1 (**Supplementary Table 5**), indicating well-controlled genomic inflation. Using the UI-ICD definitions, the liability-scale single nucleotide polymorphism (SNP) heritability, estimated using LDSC regression, ranged between 7.26% (UUI) and 11.85% (SUI) in females (assuming population prevalences of 5.36% and 4.40%, respectively) and between 2.88% (AUI) and 12.37% (MUI) in males (given population prevalences of 2.56% and 0.08%, respectively (**Supplementary Table 5; Supplementary Table 6**). Using UI-srq definitions, heritability was of 16.55% for SUI and of 14.16% (given a population prevalence of 29.86% and 27.85%, respectively (**Supplementary Table 5; Supplementary Text**)).

**Figure 1:**
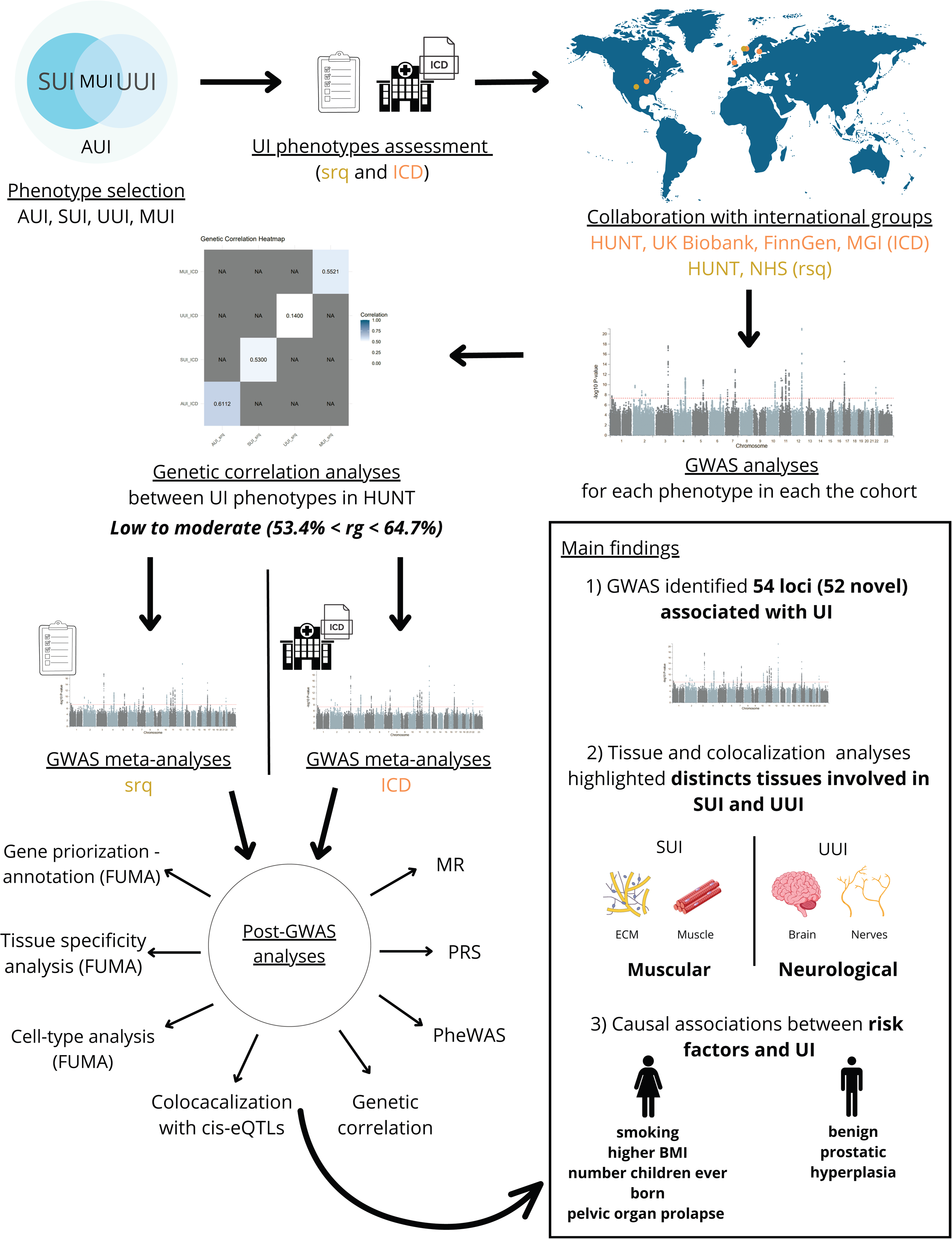
Study design and key findings. Flowchart summarizing the analytical workflow and main results. AUI; Any type of urinary incontinence. SUI; stress urinary incontinence. UUI; urge urinary incontinence. MUI; mixed urinary incontinence. ICD; International Classification of Diseases. srq; self-reported questionnaire. HUNT; Trøndelag Health Study. MGI; Michigan Genomics Initiative. MR; Mendelian Randomization. PRS; polygenic risk

The meta-analyses identified 54 independent significant loci (p-value < 5×10^−8^) associated with at least one UI phenotype, among 52 were novel (**Table 1; Supplementary Table 7-9; Supplementary Figure 1; Supplementary Figure 2**). We replicated two loci previously associated with SUI-srq in females, rs2270016 in our study in *INO80B:WBP1* (rs7607995[(8)]) and rs1448813496 near *MARCO* (rs138724718[(9)]). In total, 49 loci were exclusively found for UI-ICD definitions and four for UI-srq; the locus in *PCOLCE2* was highlighted on both. For UI-ICD definitions, we identified 37 loci in females and eight in males (**Figure 2**). No loci identified in males overlapped with those found in females or in sex-combined analyses. At each locus, the top lead variant had consistent direction across all studies, except for rs1155027 in AUI-ICD females.

**Figure 2:**
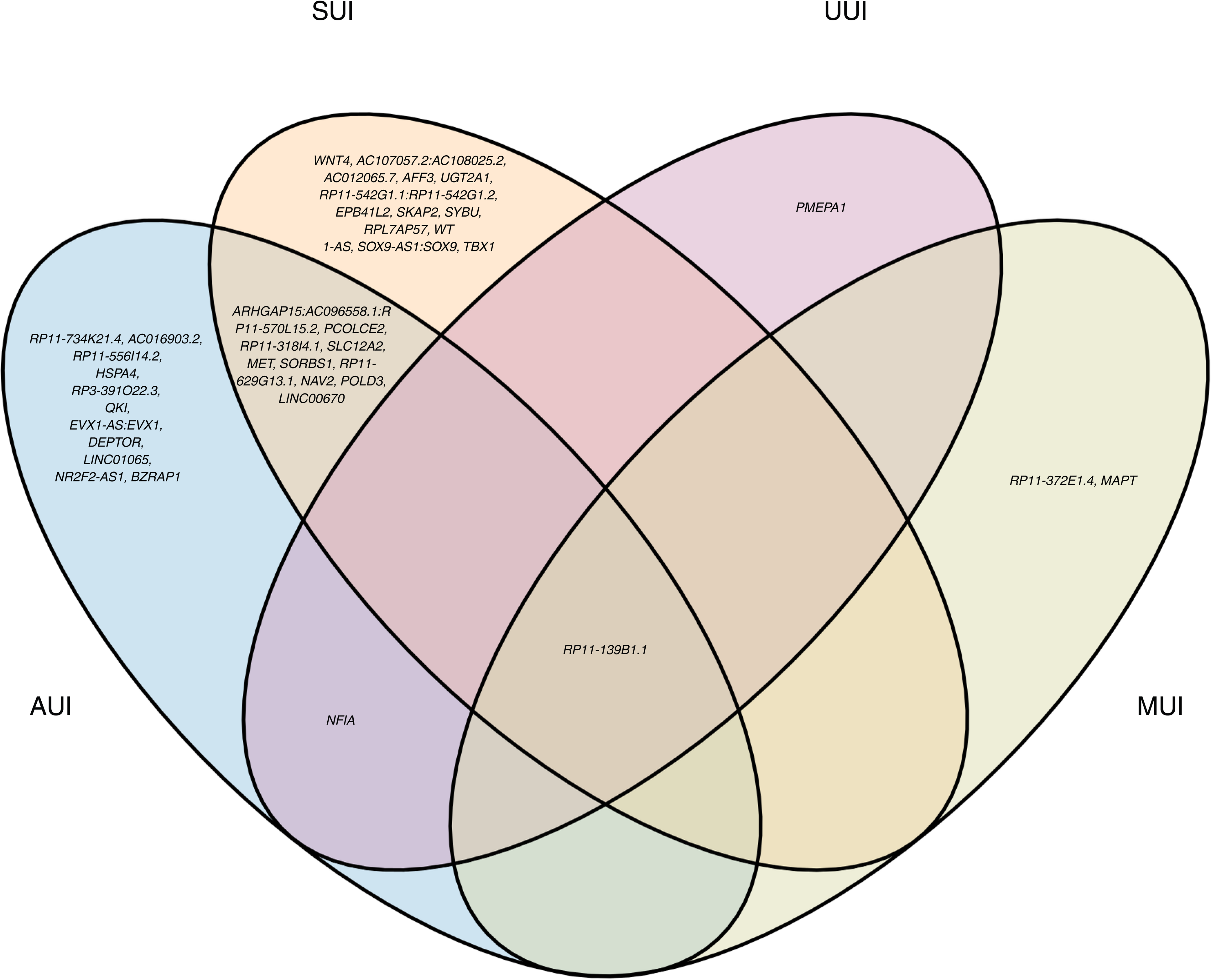
Summary of the associations for the UI phenotypes for the females (ICD based definitions). Venn diagram illustrating the small amount of genetic overlap between the main subtypes (SUI in orange, UUI in pink and MUI in yellow). The locus close to RP11-139B1.1 was shared between all the UI phenotypes. AUI; any type of urinary incontinence. SUI; stress urinary incontinence. UUI; urge urinary incontinence. MUI; mixed urinary incontinence. ICD; International Classification of Diseases.

**Table 1.**
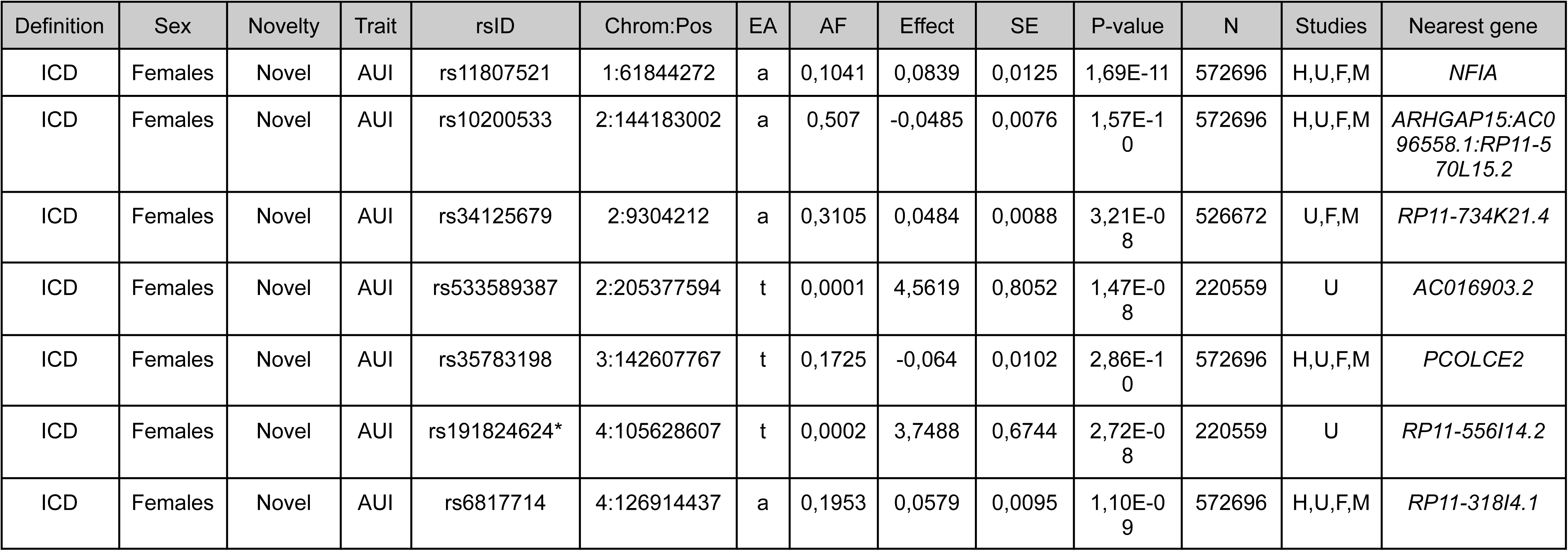
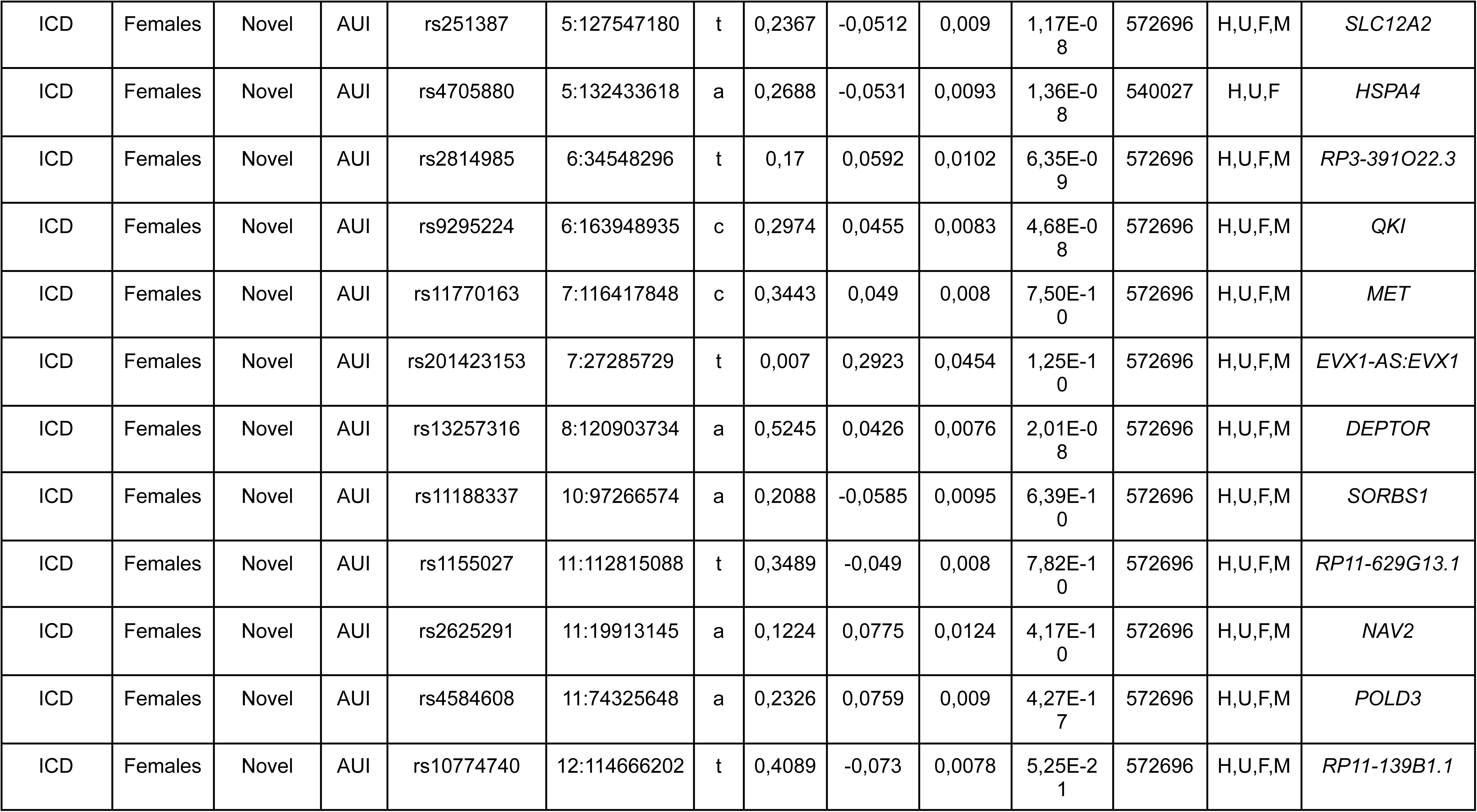
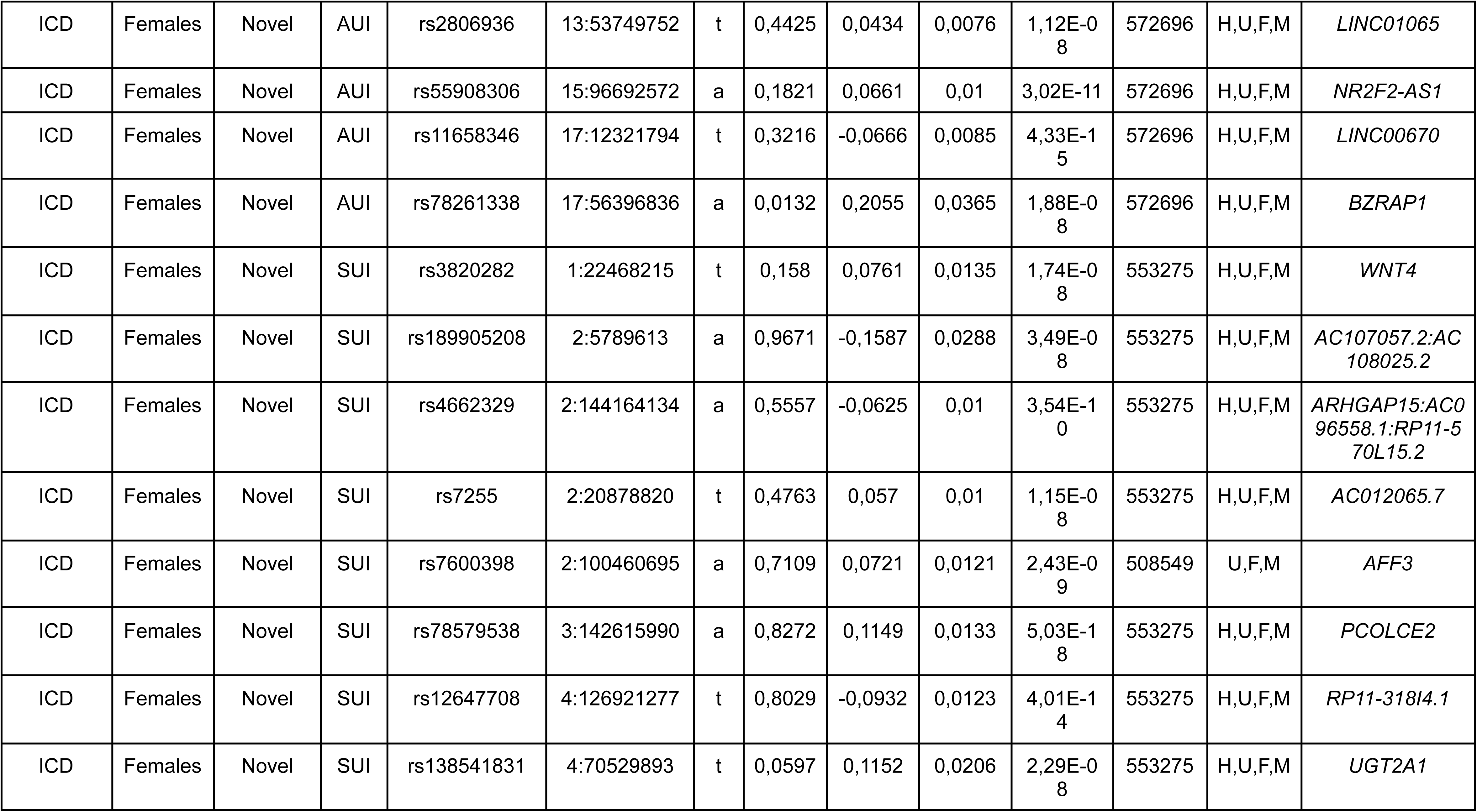
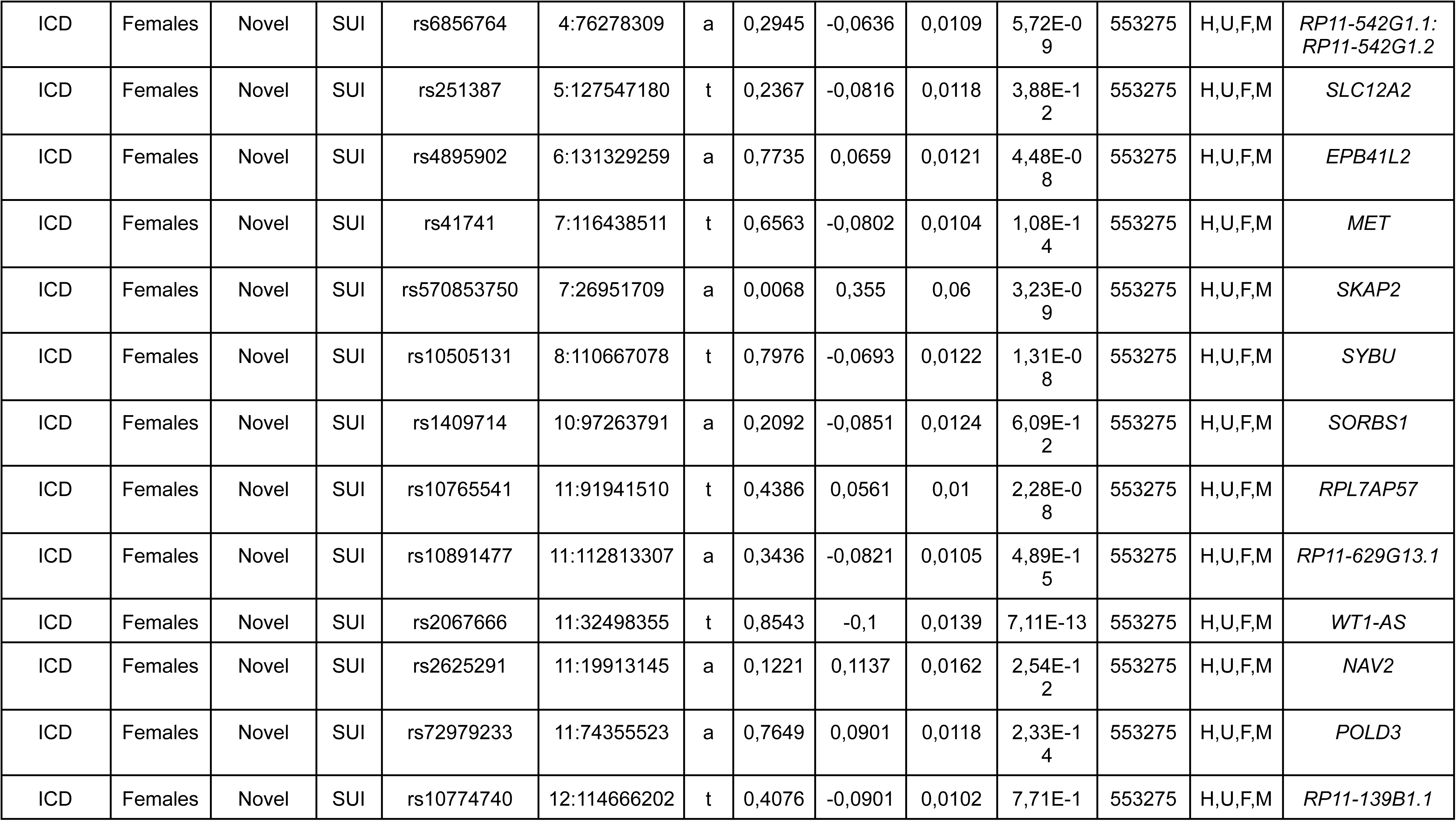
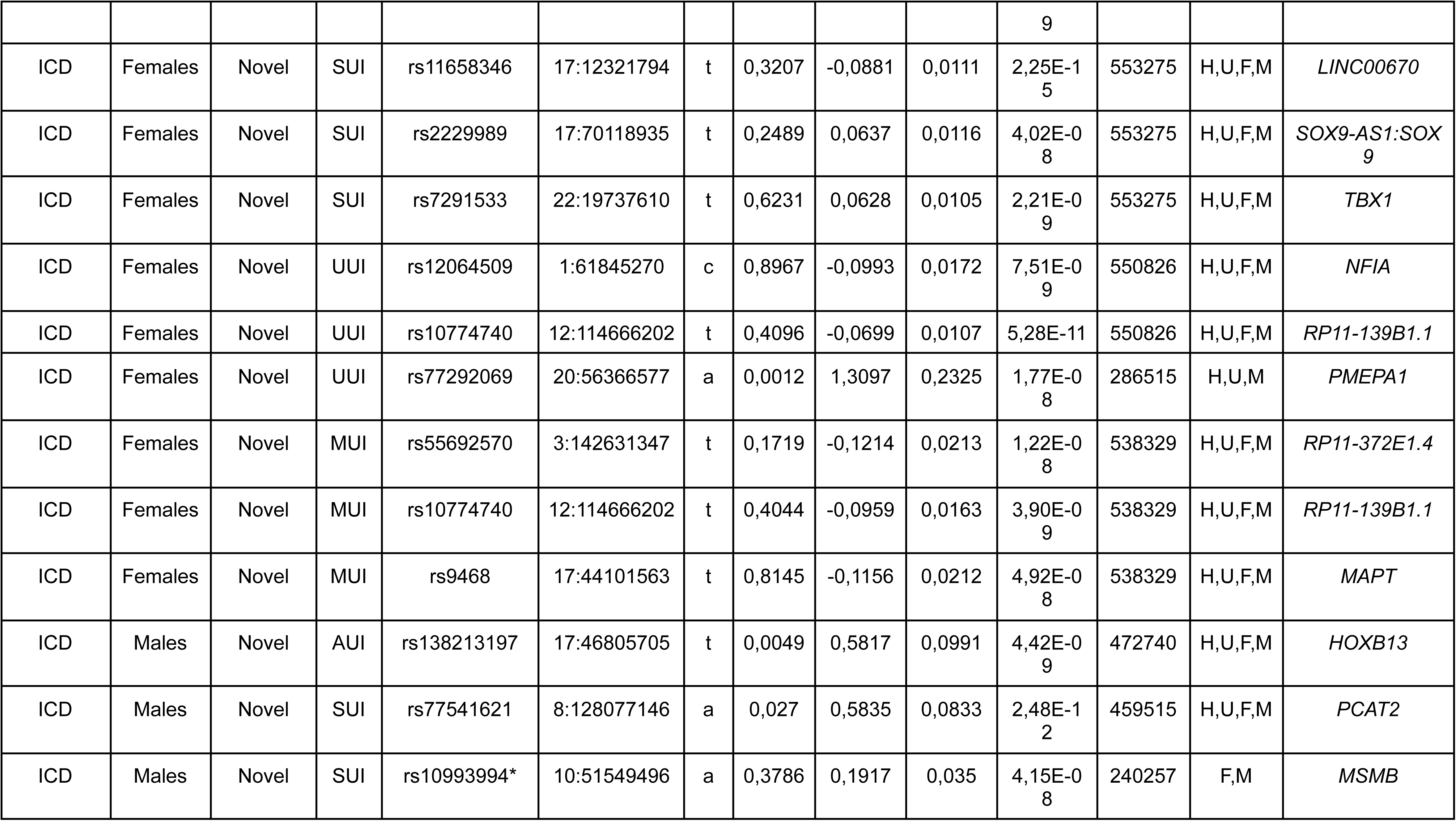
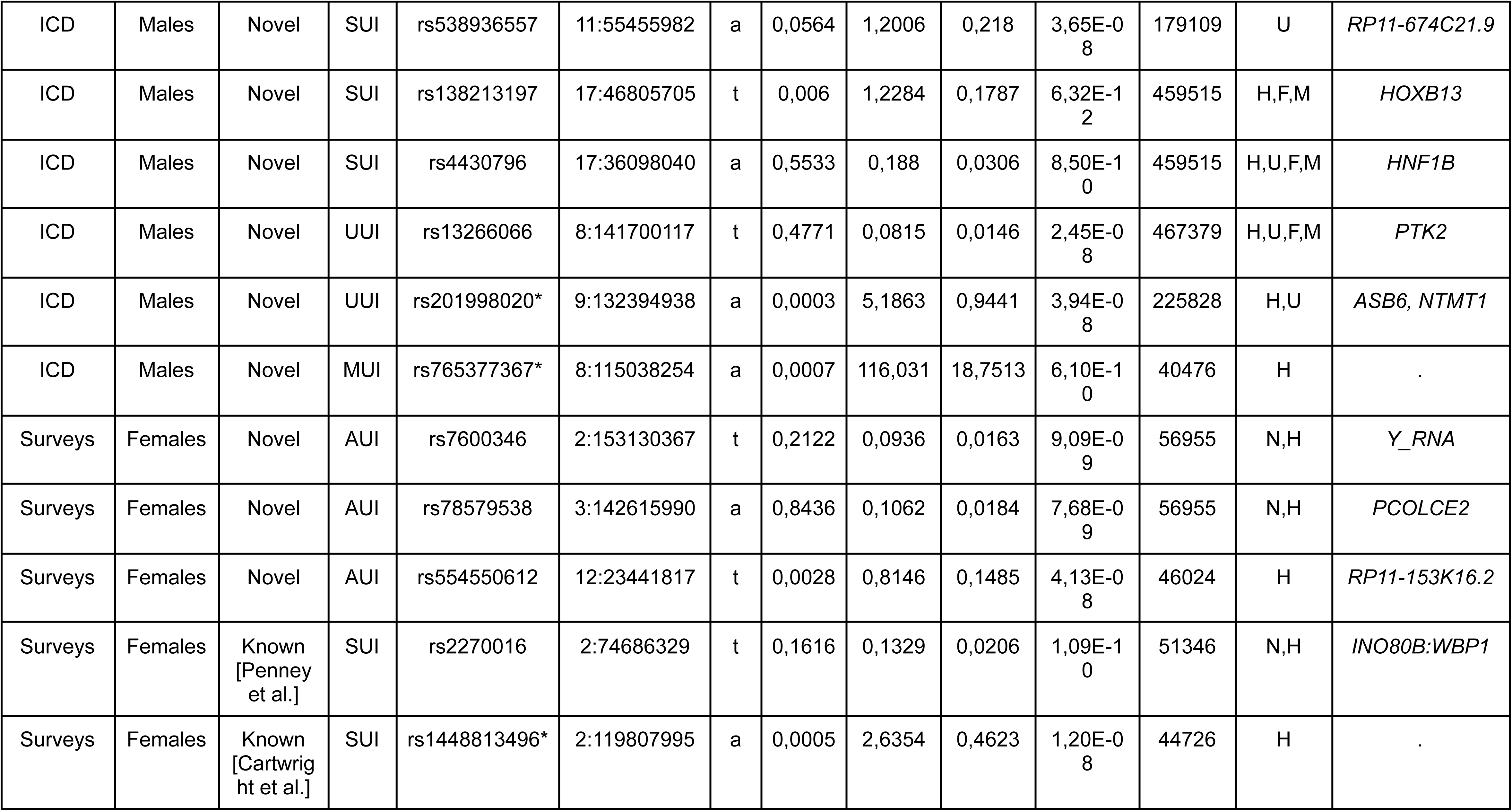
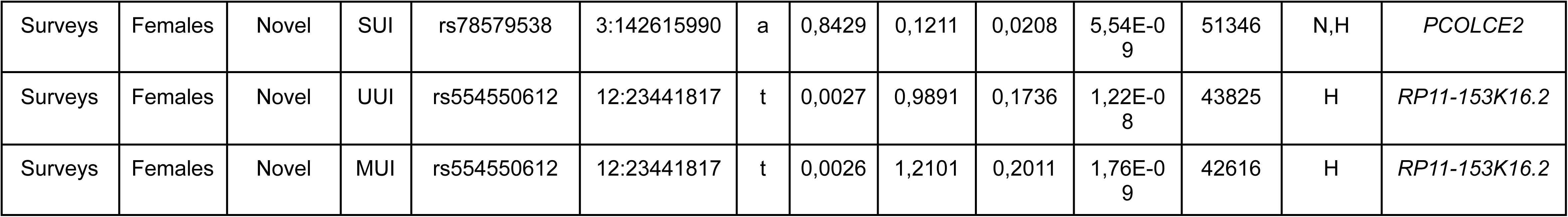
Summary of the top lead variants identified in the meta-analyses of UI phenotype. We present results for males and females for sake of clarity, results for sex-combined analyses are available in Supplementary Table 3. The loci are presented for each phenotypes: any type of urinary incontinence (AUI), stress urinary incontinence (SUI), urge urinary incontinence (UUI) and mixed urinary incontinence (MUI) for surveys and ICD phenotypes definitions (International Classification of Diseases). The variants are given with position in GRCH37. Allele frequency (AF) and effects size with standard error (SE) are given for the effect allele. Heterogeneity estimates are presented with the heterogeneity I squared (HetISq)and the heterogeneity p-value (HetPVal). N=sample size; Studies: F=FinnGen, H=HUNT, M=MGI, N=NHS, U=UK Biobank. Asterisks represents loci identified manually, filtered out by FUMA with the use of the 1000G European reference panel. For those, the Position SNP and the nearest gene were searched in VEP.

To test the robustness of our results, we performed sensitivity analyses using different ICD codes combinations and accounting for potential UI risk factors (**Supplementary text; Supplementary Table 10-13**). The beta estimates for the top lead variants obtained for AUI-ICD and UUI-ICD using alternative definitions showed consistent direction with those obtained in the main analyses (**Supplementary Table 12**). After adjustment with sex-specific risk factors of UI-ICD definitions in HUNT, beta estimates had consistent direction with those obtained in the main analyses (**Supplementary Table 13**).

### Protein-altering variants in meta-analysis loci

We identified one protein-altering top lead variant (rs138213197, non-synonymous variant in *HOXB13*) associated with AUI-ICD and SUI-ICD in males (**Supplementary Table 7**). In addition, three top lead variants were in high linkage disequilibrium (LD) (correlation r^2^ > 0.80) with 25 protein-altering variants: 1) rs2814985 (AUI-ICD in females) with one non-synonymous variant in *UHRF1BP1*, 2) rs9468 (MUI-ICD in females) with a stop-gain variant in *MAPT* and with 18 non-synonymous variants in *MAPT, CRHR1*, *SPPL2C, STH, KANSL1* and *LRRC37A2,* and 3) rs2270016 (SUI-srq) with five non-synonymous variants located in *MOGS*, *MRPL53, TTC31* and *LBX2*. The protein-altering variants mapped to genes associated with diverse traits in the GWAS catalog, spanning many biological areas (**Supplementary Table 14**).

### Prioritization of genes, pathways and tissues

To identify tissues involved in UI, we used MAGMA tissue specificity analysis to test associations between tissue specific gene expression profiles and UI-genes (identified by the MAGMA gene-based analysis). Here, we identified 11 tissues (from the Genotype-Tissue Expression [GTEx] Project version 8) in which genes with strong GWAS associations were significantly overexpressed (p-value < 9.43×10^-4^, Bonferroni correction for 53 tissue types tested) (**Supplementary Table 15**). For females, we identified uterus, tibial artery, sigmoid colon, cervix endocervix and esophagus gastroesophageal junction tissues for SUI-ICD, and cerebellar hemisphere, cerebellum, frontal cortex, hippocampus and anterior cingulate cortex tissues for UUI-ICD. In sex-combined analyses, we identified cortex and hypothalamus for UUI-ICD, in addition to the brain tissues identified in the female-specific analyses. No tissues were identified for males or for UI-srq definitions. We found seven significantly enriched gene ontology (GO) biological processes through the MAGMA gene-set analysis (p-value < 2.94×10^-6^, Bonferroni correction for the 17,023 curated gene sets and GO terms tested) (**Supplementary Table 16**).

### Cell-type analysis

To identify cell types that may play a role in SUI and UUI subtypes, we used FUMA cell type analysis to test associations between cell specific gene expression profiles and SUI/UUI-genes (identified by the MAGMA gene-based analysis). Prior to the analysis, we selected relevant cell-type datasets: bladder and muscle cell-type datasets for SUI, and hippocampus, frontal cortex and cerebellum cell-type datasets for UUI. We identified skeletal muscle satellites (from limb muscle in mouse) as the only cell type significant for SUI across all single cell RNA sequencing datasets included (p-value < 2.53×10^-4^, Bonferroni correction for the 149 cell types across all the datasets) (**Supplementary Table 17**). At dataset-level, stromal, smooth muscle and leukocyte cell types were associated with the SUI-genes in bladder tissues, additionally to mural cell type (from abdomen skeletal muscle) and tendon cell type (from pelvic diaphragm skeletal muscle). For UUI, no significant enrichment was found across all datasets (p-value < 4.31×10^-5^, Bonferroni correction for the 1159 cell types across all the datasets). However, we observed dataset-level enrichment for neurons in the hippocampus and for neuroblast in the cerebellum (uniquely in the dataset with first trimester of developmental cells).

### Colocalization

To identify shared causal variants for UI and the expression of genes nearby, and therefore strengthen the identification of candidate genes, we performed Bayesian colocalization analyses of 13 unique GWAS loci for UI and cis-expression quantitative trait loci (cis-eQTLs) in relevant tissues (**Supplementary Table 18**). We found strong evidence (posterior probability for shared causal variant (PP4) > 75%) for a shared causal variant (colocalization) of GWAS loci for UI-ICD definitions in females and eQTLs for *HSPA4* (muscle)*, MET* (tibial artery, esophagus muscularis, sigmoid colon), *DEPTOR* (brain spinal cord, esophagus mucosa, sigmoid colon), *CHRDL2* (putamen), *PCOLCE2* (fibroblast), *NCAM1* (tibial artery). For males we found *PTK2* (several brain tissues) and for sex-combined *EEFSEC* (tibial nerve). We also identified strong evidence for colocalization of GWAS loci for UI-srq in females and eQTLs for *STAM2* (muscle). Colocalization in bladder was weak using ±500 kb around the lead variant but increased markedly when restricted to a conservative ±50 kb window and the PP4 was close from 75%, suggesting the shared signal is localized near the lead variant for *LRRC37A*, *LRRC37A*2 and *KANSL1-AS1*.

### Genetic correlation of UI phenotypes

To quantify the extent of shared heritable influences between SUI and UUI subtypes, we used LDSC regression and estimated pair-wise genetic correlations (*r*_g_) of 0.86 in females (p-value= 5.70×10^-166^) and 0.58 in males (p=3.60×10^-3^) using UI-ICD definitions, and of 0.60 in females (p-value= 3.80×10^-32^) using UI-srq (**Supplementary table 19**). The pair-wise genetic correlations for SUI-ICD between females and males was of 0.17 (p-value=0.1275), and of 0.44 for UUI-ICD between females and males (p-value=7.56×10^-6^).

We also conducted pair-wise genetic correlations of UI-ICD definitions with 2,871 phenotypes from different studies publicly available (18). This revealed up to 644 phenotypes with significant genetic overlaps with the four UI types in females and 25 in males (p-value < 1.74×10^-5^, Bonferroni correction for the 2,871 phenotypes) (**Supplementary Table 20; Figure 3**). After excluding phenotypes using ICD codes related to UI or bladder disorders and for females, SUI was most strongly genetically correlated with genital prolapse (*r*_g_=0.58, p-value=1.20×10^-19^), UUI with weight gain in worst depression episode (*r*_g_=0.65, p-value=2.42×10^-8^) and MUI with reason of smoking cessation due to illness or ill health (*r*_g_=0.64, p-value=1.36×10^-5^). UI subtypes were also positively correlated (*r*_g_ > 0.3) with muscle related disorders, joint diseases, digestive related disorders, skin and subcutaneous diseases (for UUI only), circulatory system diseases (for UUI and MUI) and teeth related disorders (higher genetic correlations with UUI and MUI subtypes than with SUI). Female reproductive traits showed negative correlation with SUI and UUI. In males, UUI was most strongly correlated with hyperplasia of prostate (*r*_g_ = 0.78, p-value=1.80×10^−13^) and positive correlations (*r*_g_ > 0.29) were found also with urine retention and prostate cancer. For both sexes, UUI was positively correlated with frequency of urination and polyuria, and urinary tract infection (UTI).

**Figure 3:**
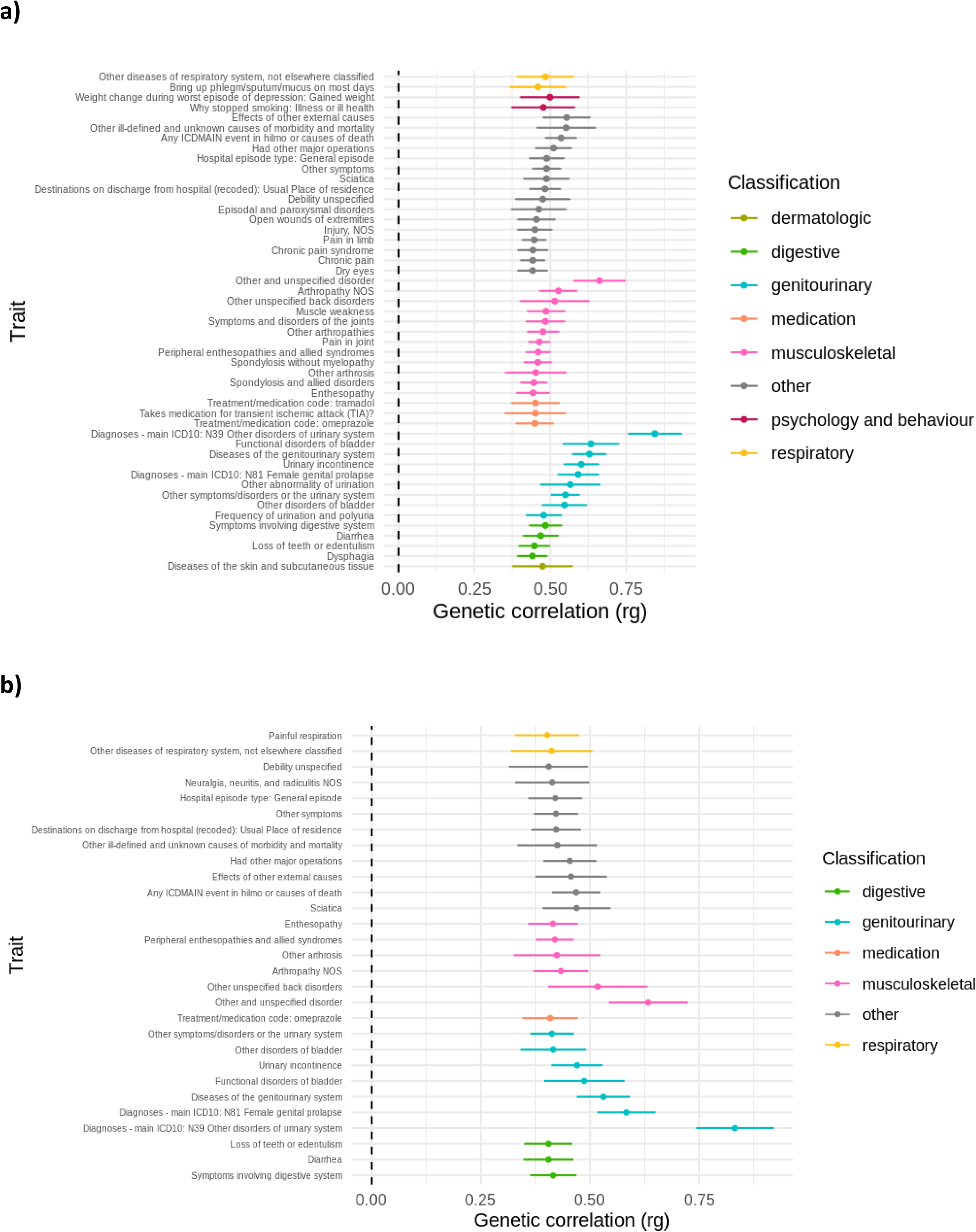
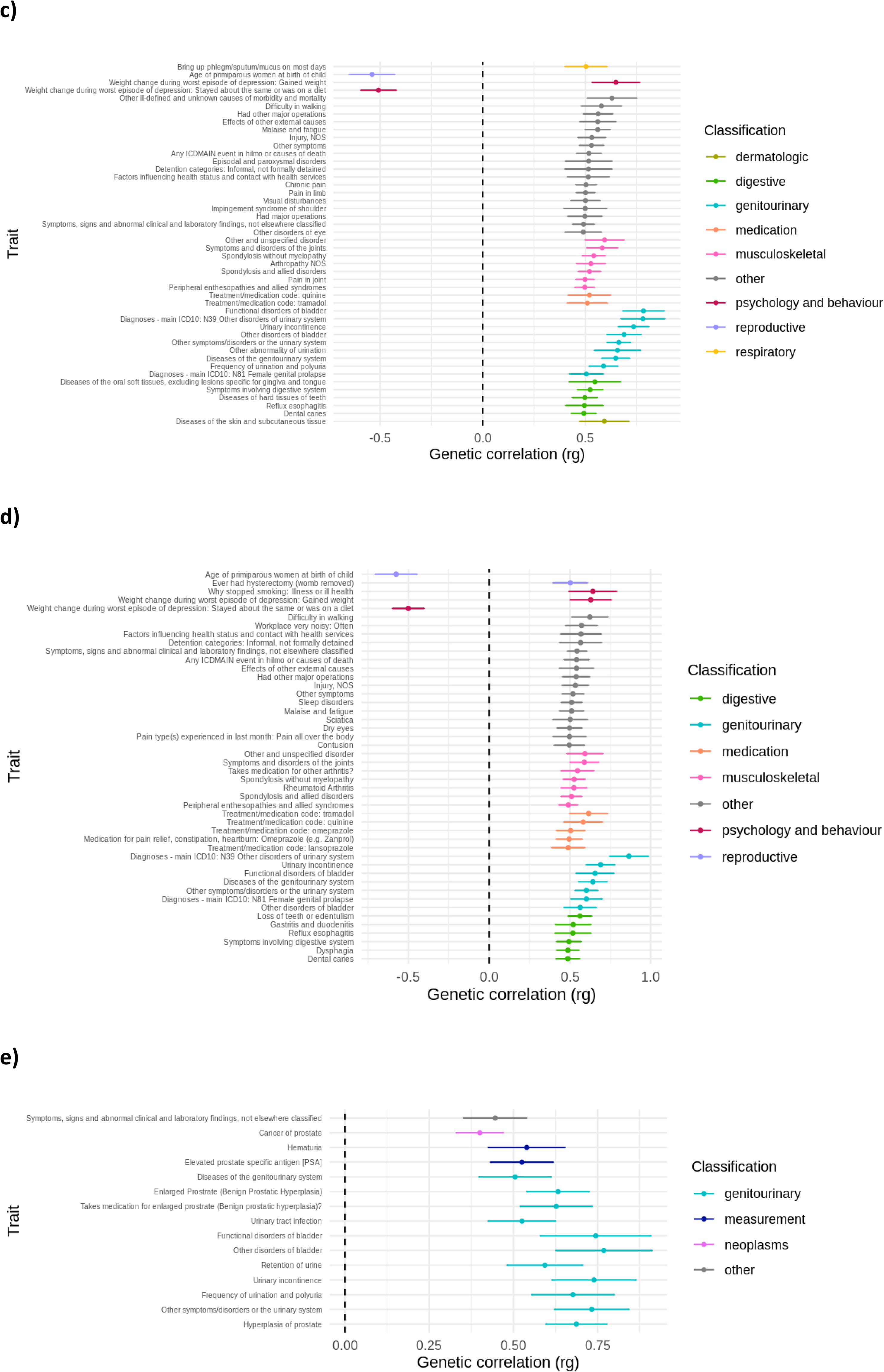
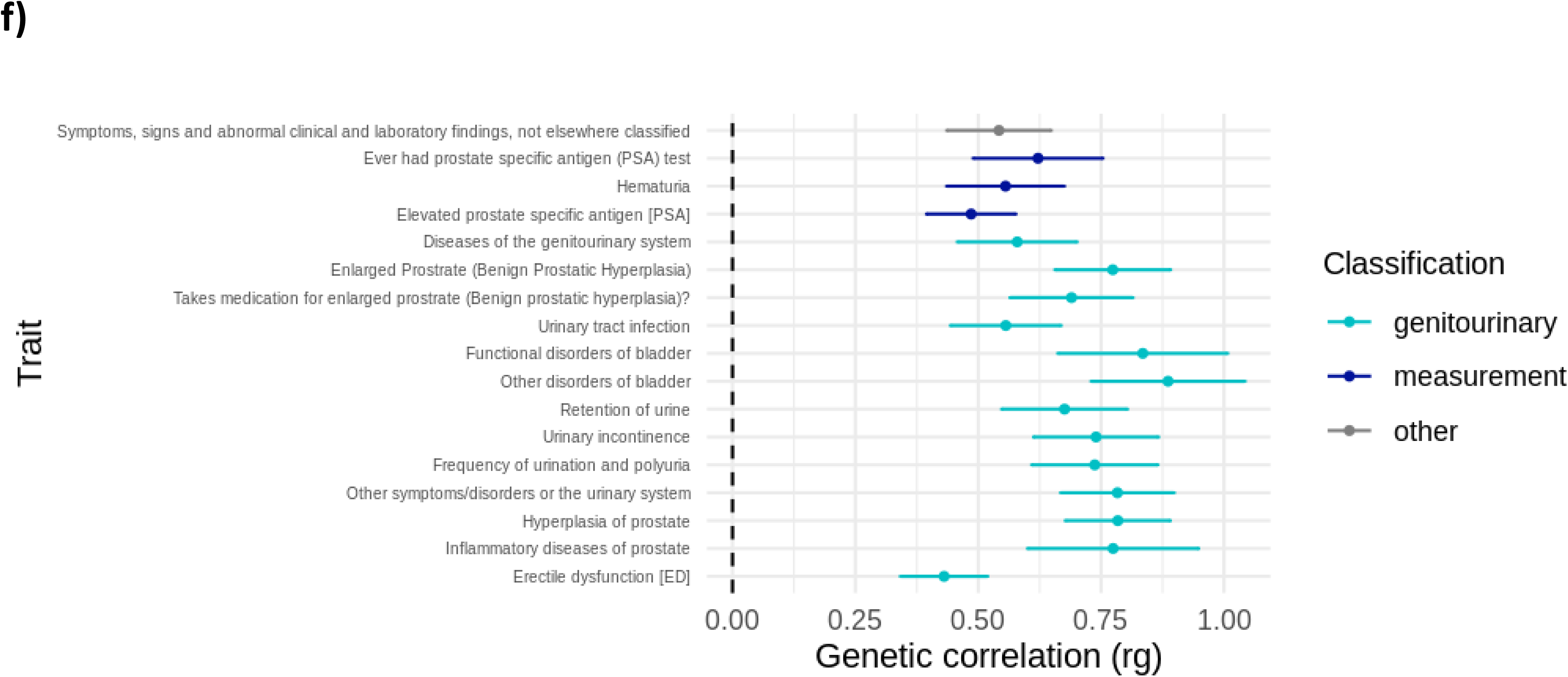
Genetic correlation analyses. Genome-wide genetic correlation of UI GWAS meta-analysis summary statistics (ICD codes) with 2,871 phenotypes. We used a Bonferroni correction for the number of tests (0.05/ 2871=1.74 × 10^−5^) and derived genetic correlation estimates (circles). Summary statistics of the phenotypes are available on the Complex Traits Genetics Virtual Lab server and come from many sources. Significant genetic correlations (rg>0.4) are plotted; we selected the first 50 largest associations if over 50 phenotypes were identified, we grouped those phenotypes by categories, corresponding to a color (under classification); a) genetic correlation associations for any type of urinary incontinence in females, b) genetic correlation associations for stress urinary incontinence in females, c) genetic correlation associations for urge urinary incontinence in females, d) genetic correlation associations for mixed urinary incontinence in females, e) genetic correlation associations for any type of urinary incontinence in males, f) genetic correlation associations for urge urinary incontinence in males

### Phenome-wide associations studies

To investigate potential pleiotropy of the top lead variants for UI with other phenotypes, we performed PheWAS using 1523 publicly available phenotypes (biomarkers, phecodes and continuous traits) from the UK Biobank. We identified 61 UI-variants as significantly associated (p-value < 4.50×10^-7^, Bonferroni correction for 1523 phenotypes and 73 variants) with at least one trait in UK Biobank (**Supplementary Table 21-23, Figure 4**). For females, we identified 18 biomarkers, 19 phecodes and 157 continuous traits associated with 26 variants. Most notably, these associations spanned the genitourinary system (mainly prolapses), the circulatory system, kidney function, metabolic traits (BMI, LDL- and HDL-cholesterol), reproductive traits (age at hysterectomy, lifetime number of sexual partners, age at first sexual intercourse, age at menarche, birth weight of first child), chronic pain, socio-economic status, smoking status and alcohol consumption. For males, we identified three biomarkers, five phecodes and 15 continuous traits associated with four variants, mainly prostate related traits (prostate cancer, hyperplasia), diabetes, kidney function, BMI related traits, circulatory system and chronic pain.

**Figure 4:**
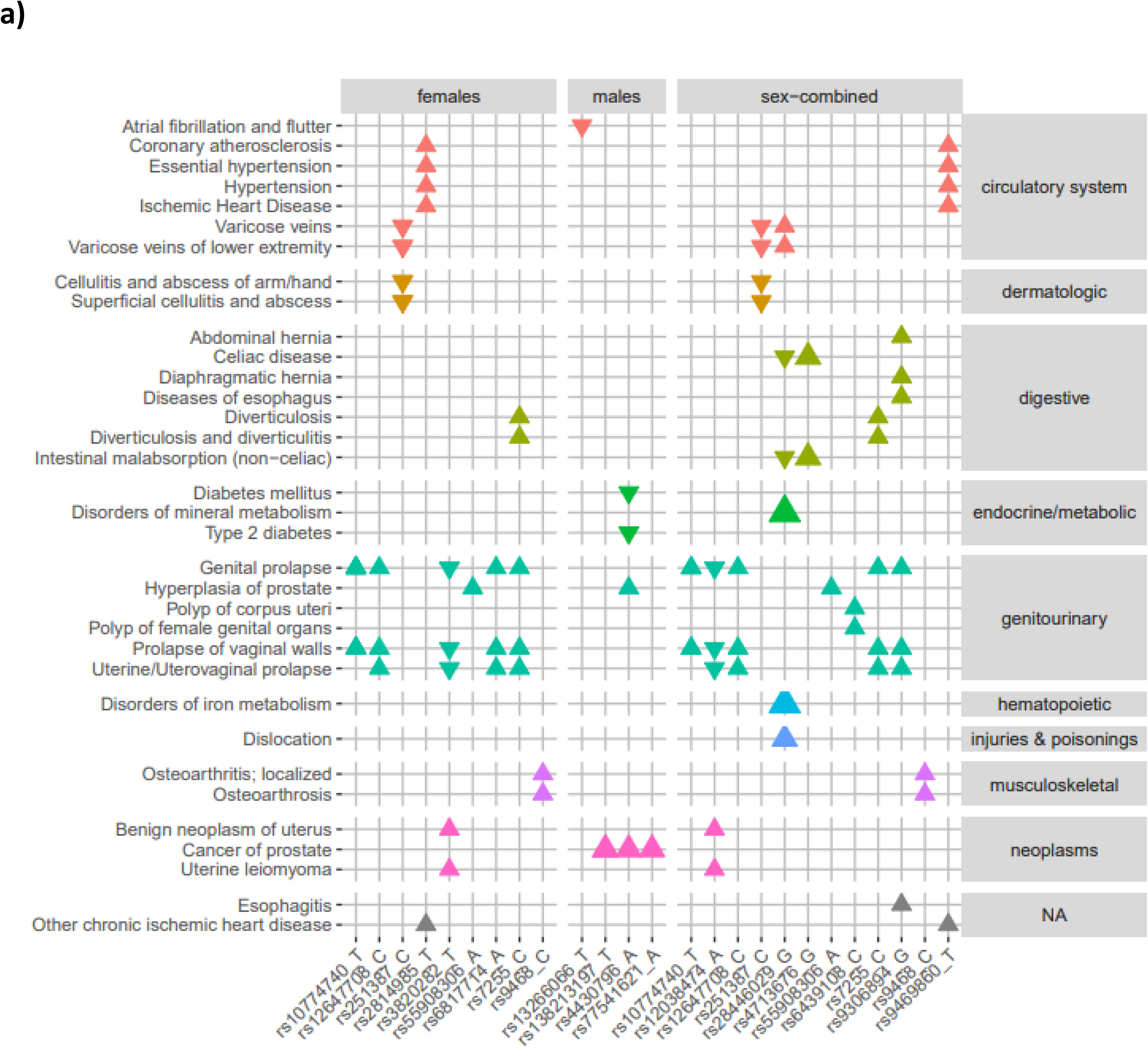
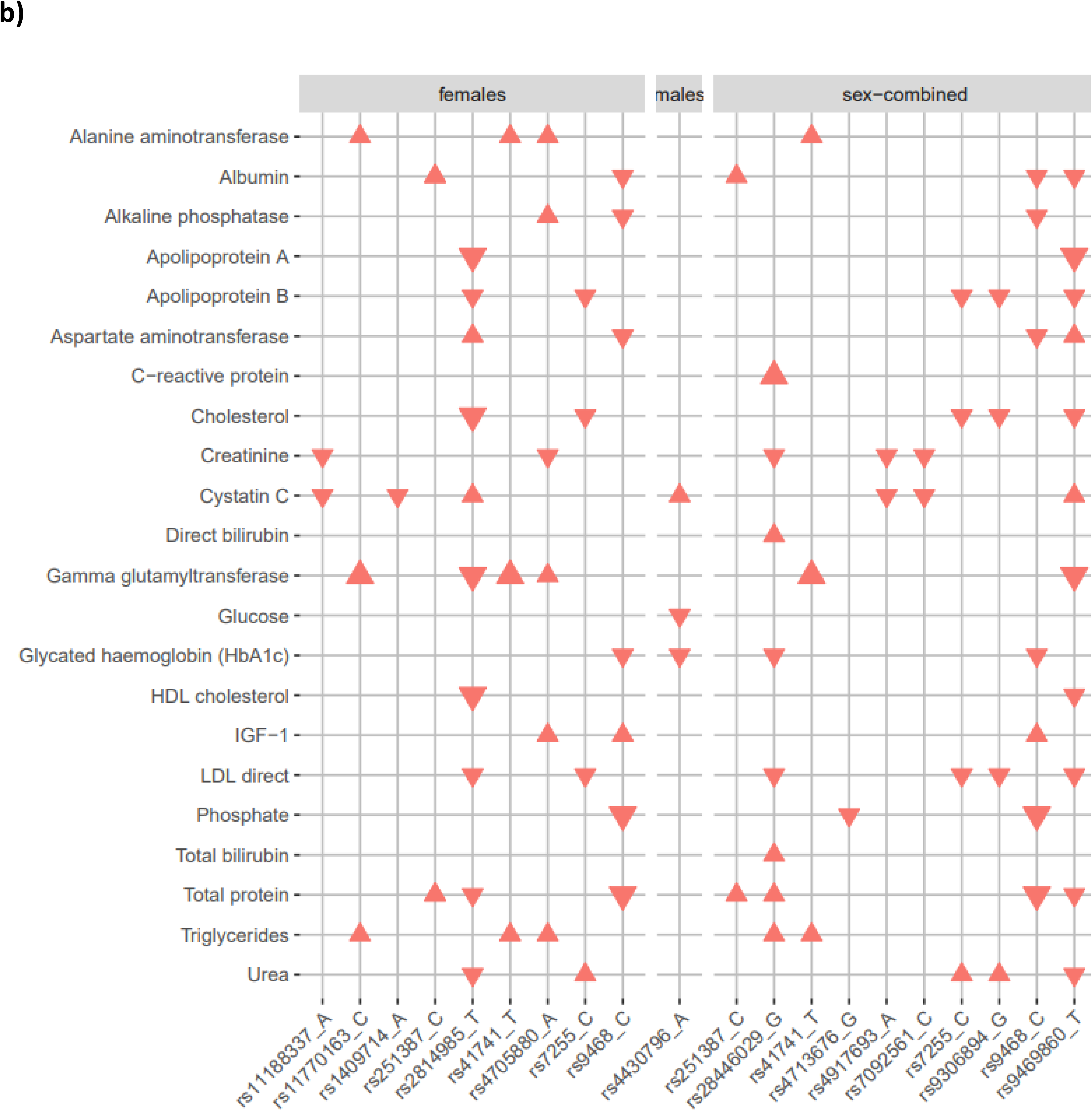
Phenome-wide associations between meta-analysis top lead variants with a) phecodes and b) biomarkers from the UK Biobank. Triangles represent statistically significant (p-value<4.5×10^-7) associations between a UI top lead variant identified by FUMA (x-axis, grouped by UI findings (females, males, sex-combined)) and a) phecode outcome from the UK Biobank (y-axis, groups by categories and colors) and b) biomarkers. Larger triangles represent strongest associations (lower p-values). The direction indicates if effect estimates (beta) directions were the same (up) or opposite (down) as the association with UI.

### UI drugs and gene target

We queried DrugBank to determine whether genes identified in our study are known targets of existing drugs for UI. Among the 13 drugs identified in DrugBank (one for SUI and 12 for UUI or overactive bladder syndrome), none were targeting a protein that is coded by a gene we identified through our meta-analysis (**Supplementary table 24**).

### Polygenic risk scores for urinary incontinence

To evaluate the potential predictive effect of PRSs in UI, we constructed PRSs using pruning and thresholding (P+T) and PRS-CS (continuous shrinkage) methods and GWAS meta-analysis summary statistics from UI-ICD definitions. The PRS explained up to 2.0% of the variation in UI in females (SUI, PRS from PRS-CS). The variance explained for males was 0.2% (SUI, PRS from PRS-CS) (**Supplementary Table 25, Supplementary Table 26**). Using PRS-CS, the prevalence in the first decile was 5.45% for SUI and 5.60% for UUI compared to 15.49% and 11.38%, respectively, in the tenth decile (**Figure 5**). In females, one standard deviation (SD) increase of the PRS was associated with an increased SUI risk of 23% (P+T) and 40% (PRS-CS), while in males it was of 7% (P+T) and 16% (PRS-CS).

**Figure 5:**
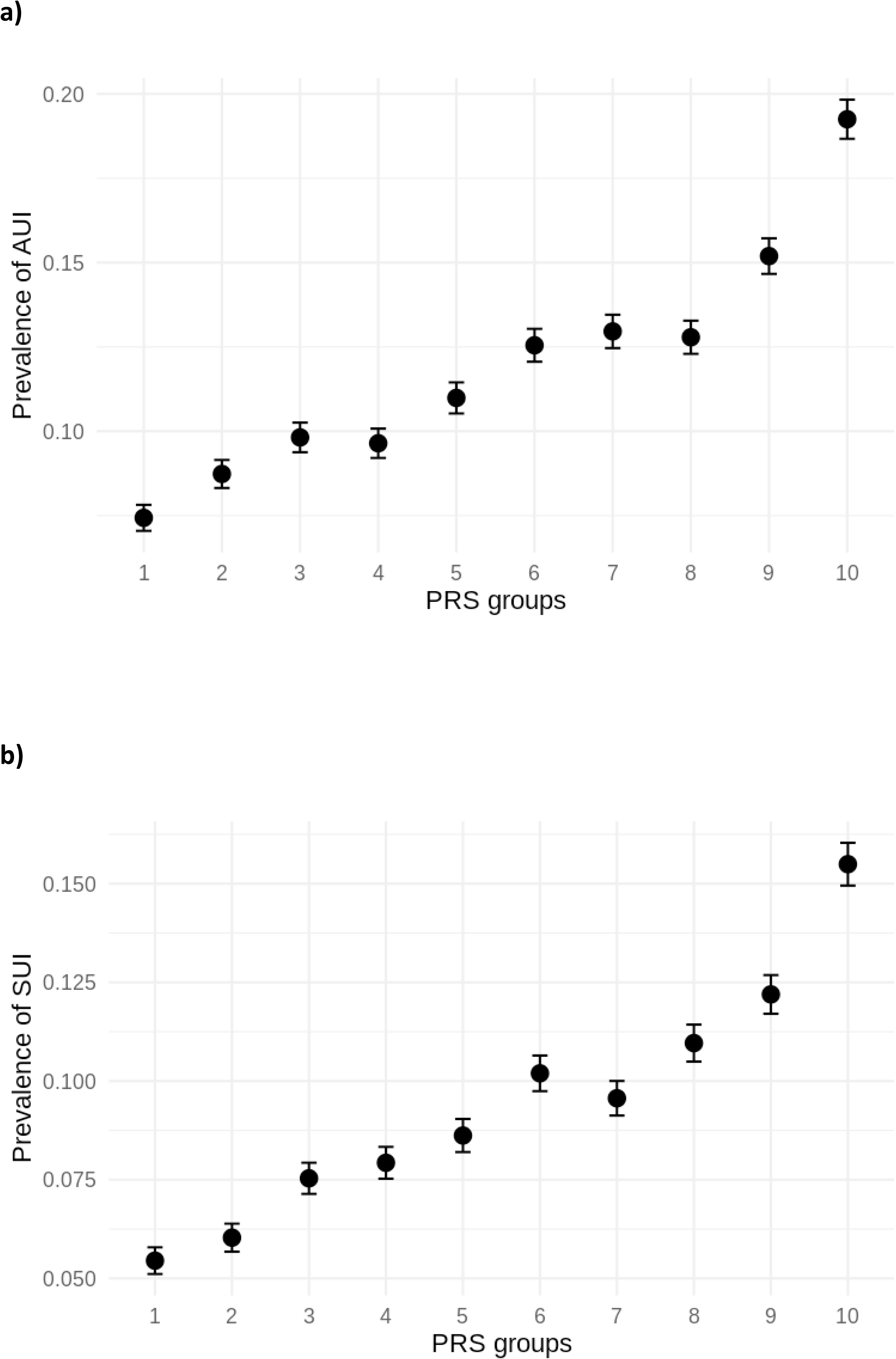
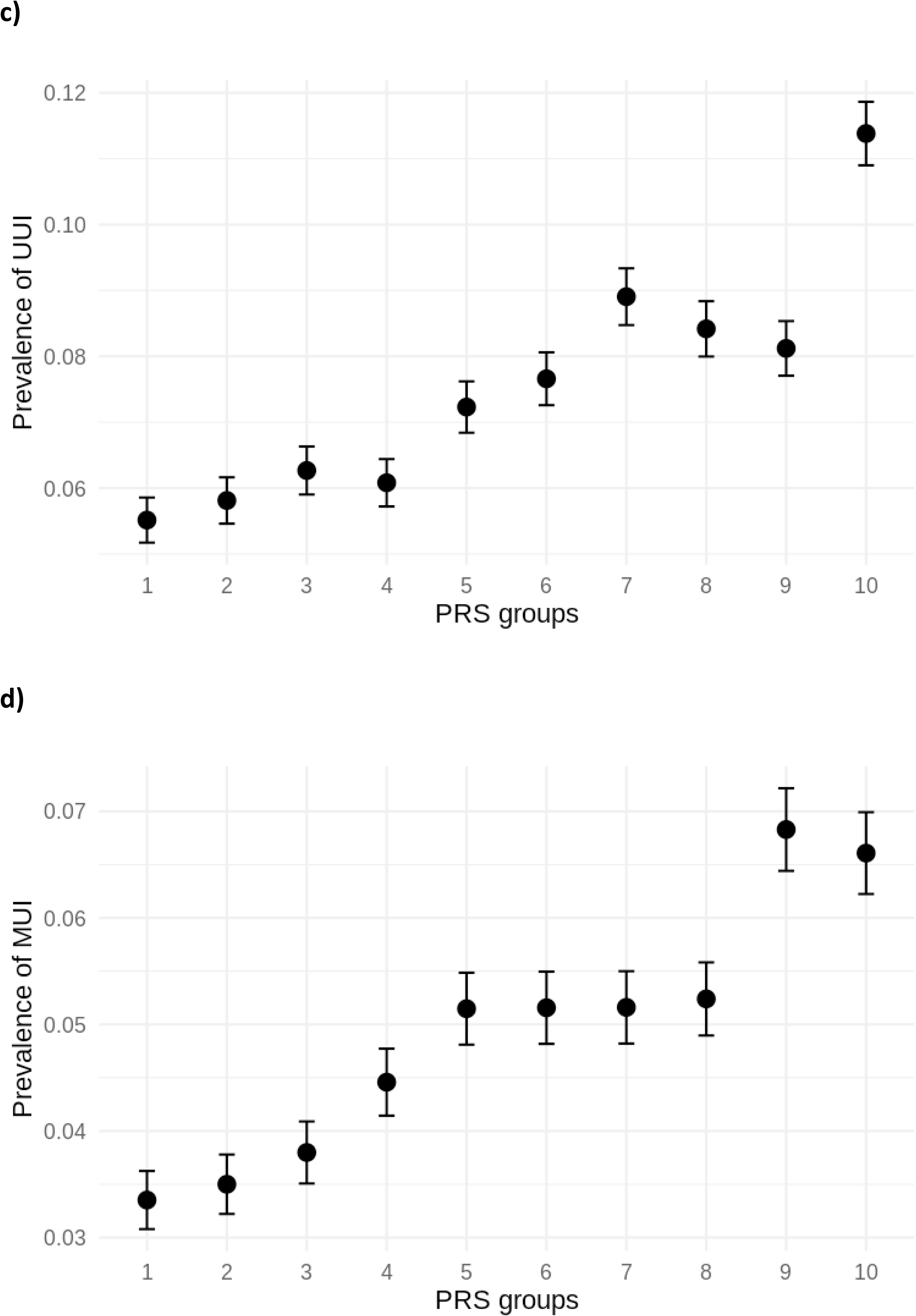
Prevalence of UI by PRS groups for females using ICD code-based definitions. a) Prevalence of any type of urinary incontinence (AUI) is presented by PRS groups using PRS-cs tested in HUNT, b) Prevalence of stress urinary incontinence (SUI) is presented by PRS groups using PRS-cs tested in HUNT, c) Prevalence of urge urinary incontinence (UUI) is presented by PRS groups using PRS-cs tested in HUNT, d) Prevalence of mixed urinary incontinence (MUI) is presented by PRS groups using PRS-cs tested in HUNT.

### Mendelian Randomization

We performed two-sample MR to investigate the causal effect of six risk factors on UI: smoking status, BMI, alcohol consumption and specifically for females, PoP and parity, and for males, BPH. In the sex-combined analysis we found evidence of a positive effect of smoking status and BMI on UI. We estimated an increased risk for UI in smokers compared to non-smokers (increased risk of 7-19% depending on UI phenotype) and per SD increase in BMI (14-36%), with consistent direction across UI phenotypes (**Supplementary Table 27**). For females, we found strong evidence for a causal effect of higher parity and PoP on UI. We estimated an increased risk for UI for each additional child ever born (82-128%) and among women experiencing PoP (12-24%), with consistent direction across UI phenotypes. For males, we found strong evidence of a causal effect of BPH on UI (2-34%), with effect size estimates consistent direction across UI phenotypes. However, the analyses for alcohol consumption showed less consistent results across UI phenotypes. Estimates from sensitivity analyses generally aligned with the inverse-variance weighted (IVW) results, although MR-Egger showed some attenuation for smoking status and BMI. Sex-stratified analyses results were consistent with sex-combined analysis results for females but not for males (IVW results) and, MR-Egger intercepts for BMI in females were close to 1 (p-value < 0.5) (**Supplementary Table 28**). Using Steiger filtering, rs4944936 explained more variance in AUI and SUI than in PoP. However, estimates remained in the same direction after excluding this SNP (**Supplementary Table 29**). All instruments had an F-statistic >10, indicating sufficient strength to minimize weak instrument bias. The genetic instruments explained between 0.2% (parity) and 5.8% (BPH) of the variance in the exposures (**Supplementary Table 30**). Except smoking status in males, all exposures showed instrument heterogeneity (Q-statistic test p-value < 0.05).

## Discussion

We performed the largest GWAS meta-analysis of UI to date and identified 54 genetic loci (52 novel) associated with at least one UI type. This replicated two previously identified loci (*INO80B:WBP1; MARCO*) for SUI in females. Analyses revealed tissue-specific involvement for subtypes of UI, with enrichment of SUI-associated genes in muscle and connective tissues, and enrichment of UUI-associated genes in neural tissues. These results were supported by key roles of SUI-prioritized genes in the extracellular matrix (ECM) structure (*PCOLCE2* and *NCAM1*) and proper muscle function (*HSPA4* and *DEPTOR*). Findings from the PheWAS and genetic correlation analyses aligned with each other, supporting the involvement of genetics in UI pathology through diverse tissue structures including connective tissues, and pathways including inflammation, immune system, and aging. Further, we confirmed the causal effects of smoking status, higher BMI, higher parity, PoP on UI in females and BPH on UI in males.

We replicated two loci out of the 12 previously reported (p-value<5×10^-8^) (8–10). The *INO80B:WBP1* locus was previously associated with SUI in models adjusting for BMI, parity, and diabetes, whereas we identified it in our main analysis (models non-adjusted for clinical factors) (8). Among the six GWAS of UI previously published, only three reported loci at the genome-wide significance threshold of p-value<5×10^-8^. This could be explained by 1) heterogeneous phenotype definitions from self-reported questionnaires and ICD codes, which may not be comparable, 2) a focus of females-specific GWAS and, 3) the limited sample size (n<26,000) of previous studies. We did not replicate genes from candidate gene studies (7,19).

We identified three top lead variants in high LD with protein-altering variants: 1) rs2814985/rs9469860 were mapped to *UHRF1BP1* and *SNRPC*. Individuals with mixed connective tissue disease frequently produce autoantibodies against a protein encoded by *SNRPC* (20). Given that connective tissue integrity is essential for bladder support, its dysfunction may contribute to UI. 2) rs9468 was in high LD with protein-altering variants in genes involved in tauopathy, dementia and Alzheimer’s disease (*MAPT* and *STH)*. 3) rs2270016 was in high LD with a protein-altering variant in *LBX2, MOGS, MRPL53;* these genes have been suggested to influence SUI with unclear described roles (13). As replicated in our PheWAS, the protein-altering variant rs138213197 in *HOXB13* is known to be associated with prostate cancer (21,22); a likely comorbidity causing UI.

In females, our findings suggest sex-specific tissues to be involved in UI subtypes: smooth muscles, connective tissues and mucosa tissues for SUI, and brain related tissues for UUI. Smooth muscle constitutes the detrusor muscle of the bladder, allowing the bladder to relax during filling phase and to contract during the voiding phase. The smooth muscles also constitute urethral sphincter which regulates its opening to prevent urine leakage. This is done in coordination with connective tissues and pelvic floor muscles. Connective tissues maintain the structural integrity of organs and, in the context of urinary continence, provide structural support to the urethra and bladder through the ECM. Within the ECM, fibroblasts produce collagen and other structural proteins, providing tissue elasticity and strength and the bladder mucosa is composed of ECM. Disruption of any of these components may contribute to UI.

These findings are supported by the role of the SUI-prioritized genes in the ECM (colocalization in fibroblasts and tibial arteries, a tissue composed of connective tissues)): *PCOLCE2* by playing a role in collagen formation (23,24) and *NCAM1* by encoding a cell surface glycoprotein that mediates cell-cell and cell-ECM matrix interactions (25,26). Secondly, SUI-prioritized genes are known to play a role in muscle cell structure, support and contraction: *HSPA4* (colocalization in muscles) is known to support protein cellular homeostasis maintenance and was upregulated during muscle regeneration (27), while *DEPTOR* (colocalization in gastrointestinal tissues) regulates myocyte protein metabolism, and its reduced expression can improve muscle atrophy (28). Thirdly, smooth muscle cells, satellite cells, and tendon cells were cell types associated with SUI-genes. While the smooth muscle cells form the detrusor muscle and enable the bladder to relax (filling phase) and to contract (voiding phase) (29), the satellite cells contribute to tissue regeneration (30) and the tendon cells form the tendinous arch, a dense fibrous connective tissue anchoring muscles to the pelvic bone, supporting continence (3,31).

The findings for UUI tissues in females are firstly supported by the role in neurological disorders of UUI-prioritized genes, that suggest neural pathways*: PTK2* (colocalization in diverse brain tissues) has been associated with tau-induced neurotoxicity regulation in neurodegenerative diseases and *EEFSEC* (colocalization in tibial nerve) with neurodevelopmental disorders with progressive neurodegeneration. Although these findings could suggest potential comorbidities, the absence of associations with neurodegenerative diseases in genetic correlations and PheWAS analyses reinforces that the prioritization of these genes may not be due to such comorbidities. Secondly, brain tissues involvement may reflect neurogenic detrusor overactivity and can lead to UUI due to a disrupted bladder-central nervous system coordination after surgery, injury or disease. Our findings align with neuromodulation, current therapies for UUI which target bladder, nerves, brain signaling pathways. The main enrichment analysis included the *NFIA* locus, previously associated with UI (32); however excluding this locus yielded similar tissue associations, thereby indicating that brain enrichment was not only driven by this locus. Thirdly, the cell-type analysis identified neuroblast cells from the cerebellum and neurons from the hippocampus associated with UUI-genes. The cerebellum coordinate the detrusor contraction, control the pelvic floor in the micturition reflex and activate during the filling phase (33,34).

The findings from the genetic correlations and the PheWAS aligned with each other, supporting the involvement of genetics in UI through ECM and connective tissues structures. In females, we found genetic correlations of UI with ECM-related and connective-related disorders such as PoP, chronic periodontitis, osteoarthritis (involve ECM but not connective tissues) and myositis. Similarly, seven of nine prolapse-associated SNPs had same directions with UI, and rs9306894-G was associated with esophagus disorders, suggesting the involvement of connective tissues. We also observed the association of rs9468-C with UI and osteoarthrosis with concordant effect directions. Additionally, the genetic correlations of PoP with UI could reflect the involvement of smooth muscles and the correlation of musculoskeletal disorders (e.g. myalgia, myositis and muscle weakness) with UI could reflect the involvement of skeletal tissues. Our findings also support the involvement of genetics in UI through several pathways, mainly those related to inflammation and immune system. We found correlations of UI with many inflammatory conditions such as gingival and periodontal diseases, chronic periodontitis and rheumatoid arthritis. Our findings suggest that the previous reported association between overactive bladder and tooth loss due to chronic periodontitis (35) may be driven by shared genetic factors, indicating common shared biological pathways. We also highlighted moderate correlations between UTI and UUI. Finally, the association of rs9468-C with UI and osteoarthrosis supports an age-related pathway, given that osteoarthrosis has a higher prevalence in older adults.

Current pharmacotherapeutics for UI target subtype-specific mechanisms which align with our findings, particularly for UUI. Medications for UUI target neural signaling by acting as antagonists of muscarinic receptors or β3-adrenergic receptor agonists. Antagonists of muscarinic receptors reduce parasympathetic cholinergic activity and therefore involuntary detrusor contractions, while β3-adrenergic receptor agonists enhance sympathetic signaling and promote smooth muscle detrusor relaxation during the filling phase to reduce urgency. In contrast, the only listed medication for SUI in the DrugBank (Duloxetine) acts by enhancing urethral sphincter contraction via central and peripheral neural mechanisms. While this improves functional sphincter deficit observed during intra-abdominal pressure, it does not directly target urethral muscle structure, connective tissues or ECM, which our analyses identified as central in SUI pathophysiology. Further, none of the approved drugs target proteins encoded by genes identified in our meta-analysis, highlighting a potential for genetically informed therapeutic development and repurposing.

Despite the convincing roles of the associated genes in UI pathology, the PRSs we constructed had limited predictive power. Given the range of PheWAS traits identified, other clinical comorbidities may serve as stronger predictors for UI risk.

Two-sample MR confirmed the causal effect of smoking status, higher BMI, higher parity, PoP and BPH on UI, aligning with previous observational studies (14–17). Sex-specific analysis suggested that smoking status and BMI were clear risk factors for UI in females but not in males. Our results for alcohol consumption were not consistent across all phenotypes. However, we observed a protective effect on UUI in males, as previously reported in an observational study including men (36). Through MR we highlighted causal associations between parity and UI, and through PheWAS we find genetic variation for UI to also be associated with birth weight of the first child. These findings highlight that childbirth is a critical period for women to develop UI and reinforces the importance of raising awareness to empower women to recognize symptoms, report them without stigma and seek appropriate care. It also emphasizes the need for clinicians to proactively initiate discussions about UI pre-and post-partum and manage the symptoms accordingly.

This study had several advantages and clear limitations. To date, this is the largest meta-analysis of ICD code-based definitions (>1M individuals) of UI and of self-reported UI (>56,000 individuals). The large sample size, and consistent effect directions support the generalization of our findings. However, the study was limited to individuals of European ancestry. Future replication in populations with diverse genetic ancestries is needed. UI is a broad term encompassing multiple ICD codes. Due to country-specific usage of the ICD code system, we collaborated with clinicians in Norway and the US to define the phenotypes variables. Because ICD codes may only capture severe cases of UI or patients with comorbidities; we also used self-reported data to identify moderate cases. Given these differences, we estimated the genetic correlation between self-reported and ICD code-based definitions and analyzed them separately. Population-based self-reported data was used to represent the general population while hospital-based data increased sample size and variant detection. However, this may have introduced selection bias, as hospital participants are more likely to present with severe UI, comorbidities or potentially have higher health literacy promoting health-seeking behavior. Further, our sensitivity analyses using all World Health Organization ICD codes for UUI and AUI definitions, and adjusted for potential confounders, did not show material differences to our main analyses, supporting the robustness of our results. Finally, no overlapping loci were found between sexes, suggesting distinct genetics for UI. However, this could result from limited sample size and lower prevalence in males. This was supported by the female-derived PRSs being associated with an increase of UI in males (**Supplementary Table 25; Supplementary Table 26**).

In conclusion, our study increased the number of loci associated with UI through the largest GWAS meta-analyses of hospital-based and self-reported data to date. We identified sex- and subtype-specific genetic mechanisms underlying UI and highlighted tissue-specific involvement in the two main subtypes of UI. We confirmed the causal effect of smoking, higher BMI, parity and PoP on UI in females and BPH on UI in males. Our findings enhance our understanding of UI genetics, underlying tissues, and causal factors. Finally, these results highlight the need of genetically informed therapeutic development and repurposing.

## Methods

### Cohort descriptions

#### HUNT

The Trøndelag Health Study (HUNT) is a large Norwegian longitudinal population-based study (37–39). Self-reported data, clinical measurements and biological samples have been collected over forty years: HUNT1 [1984-1986], HUNT2 [1995-1997], HUNT3 [2006-2008] and HUNT4 [2017-2019]. In total, ∼229,000 adults (≥20 years) have participated in at least one HUNT study and ∼88,000 have been genotyped (40). Participants were linked to electronic health records allowing for a longitudinal follow-up using ICD codes available at the local hospitals starting from 1987. Sample and genetic variant quality control (QC) have been reported elsewhere (40).

#### UK Biobank

The UK Biobank is a population-based database gathering self-reported health information, clinical measurements and biological samples for ∼500,000 adults living in the UK (aged 40-69 years) (41). At baseline, participants consented to link their data to electronic health-related records (National statistics, hospital information and primary care data) for follow-up. In total, 488,377 participants were genotyped (41).

#### FinnGen

FinnGen is a Finnish public-private collaborative study combining data from digital health record from Finnish health registries with genotyping data (42). The data freeze 12 includes 520,210 participants. Sample and genetic variant QC are described elsewhere (42,43).

#### MGI

The MGI is a biobank including patients receiving care at the Michigan Medicine medical center from 2012 onwards. Approximately 95,000 participants (aged ≥ 18 years) consented to link their genetic data to their electronic health records (44); therefore, in the Data Freeze 6 > 80,000 participants had genotyping data. Sample and genetic variant QC are described elsewhere (44).

#### NHS

The NHS cohort was initiated in 1976 and included >121,700 nurses (aged 30-55 years) in the NHSI. Participants answered biennial questionnaires about their health and lifestyle and provided blood and cheek cell samples (45). Similarly, the NHSII recruited 116,430 additional nurses (aged 25-42 years in 1989). Sample and genetic variant QC are described elsewhere (8,46). We used summary statistics from Penney and al. 2019 using NHS data and publicly available in the GWAS Catalog (8).

For each cohort, the details of the genome-wide genotyping, imputation and quality control methods are provided in (**Supplementary Table 31**).

### Phenotype definitions

#### Self-reported phenotype definition

##### HUNT

Answers about UI were collected in the baseline questionnaires in HUNT 2-4. The questions differed between the self-reported questionnaire, sexes and age (**Supplementary Table 32**). We combined the questions from the HUNT self-reported and constructed four phenotype definitions: any type of UI (AUI-srq), SUI (SUI-srq), UUI (UUI-srq) and MUI (MUI-srq) (Phenotype definitions and number of individuals included are presented in **Supplementary Text; Supplementary Table 3**).

##### NHS

Four definitions have previously been used: SUI, UUI, MUI and all UI. In brief, UI cases reported at least weekly UI at most questionnaires (≥4 from the 7 questionnaires in NHSI, ≥3 from the five questionnaires in NHSII). Definitions are reported elsewhere (8) and the number of individuals included in our analysis is presented in **Supplementary Table 3**.

#### ICD-code based phenotype definitions

We constructed four ICD code-based UI definitions: any UI (AUI-ICD), SUI (SUI-ICD), UUI (UUI-ICD) and MUI (MUI-ICD) by combining ICD Ninth Revision (ICD-9) and tenth Revision (ICD-10) codes. We used definitions from the WHO dictionary for ICD-10 (version 2019; https://icd.who.int/browse10/2019/en#) for HUNT, UK Biobank and FinnGen and ICD-10-CM for MGI from the National Center for Health Statistics (version October 1^st^, 2023; https://ftp.cdc.gov/pub/Health_Statistics/NCHS/Publications/ICD10CM/2024/). The phenotype definitions were constructed in collaboration with local physicians to ensure that they correctly represented the coding systems used in Norway and in the US (**Supplementary Table 33, Supplementary Text**). The number included for each definition are presented **Supplementary Table 2**.

##### Genome-wide association analyses

We performed GWAS using a generalized mixed model as implemented in SAIGE (47) version 1.03 for GWAS using HUNT data and version 0.35.8.3 for GWAS using UK Biobank data. We adjusted the analyses for age, sex, the first ten genetic principal components (PCs) and genotyping batch. We excluded variants with minor allele count (MAC) < 10 or imputation INFO score < 0.3. We inspected QQ plots, estimated the genomic inflation factor (lambda) and intercepts using LD score regression (48,49).

##### Meta-analyses

We used METAL to conduct 16 fixed-effect inverse variance weighted GWAS meta-analyses of the effect size estimates and standard errors of 1) the HUNT questionnaire-based and NHS summary statistics including up to 56,957 individuals of European ancestry (50) and 2) ICD code-based definitions from HUNT, UKBB, FinnGen and MGI including up to 1,045,436 individuals of European ancestry. Prior to the meta-analysis, we converted the genomic positions in HUNT and UKBB from GRCh37 to GRCh38 using LiftOver from UCSC (51). In addition, we applied genomic control correction based on the estimated inflation factor (lambda) for each cohort. Prior to meta-analyses, we assessed the genetic correlations between ICD code-based definitions and self-reported definitions from HUNT; the genetic correlation was low to moderate (0.53 < *r*_g_ < 0.65) and we decided to treat the phenotype definitions separately (**Supplementary Table 34**). We checked for heterogeneity using the I^2^ from METAL (acknowledging heterogeneity if p-value < 0.05/(number of top lead SNPs for the phenotype)) (**Supplementary Table 7**). Using the summary statistics from the meta-analyses, we calculated the lambda (minor allele frequency > 0.01) (**Supplementary Table 4**) using all the variants available and estimated the intercept (**Supplementary Table 5**) using the LD Score regression (48,49), restricting to the variants present in the HapMap3 SNP-list.

##### Sensitivity analysis

To check whether the identified loci were reflected by known risk factors more than UI, we performed GWAS using HUNT adjusted for known UI risk factors (**Supplementary Text**). Additionally, we performed GWASs using alternative combinations of ICD codes to investigate robustness of main analysis definitions as described in the **Supplementary Text**.

##### Genomic risk locus definition

We used FUMA v1.6.1-1.6.5 (52) (https://fuma.ctglab.nl/) to annotate and positionally map variants from the meta-analyses after converting genomic positions to build GRCh37 using the UCSC LiftOver (51). This was done because the FUMA versions used did not handle data of build GRCh38. Genomic risk loci were defined by FUMA using default parameters (maximum lead SNP p-value < 5×10^−8^, r^2^ threshold for independent significant SNPs ≥0.6; 2nd r^2^ threshold for lead SNPs ≥0.1, maximum distance of LD blocks to merge ≤ 250kb (kilobases)). The SNP with the smallest p-value in each locus is termed ‘top lead SNP’ in FUMA, a term we kept here for clarity. We used LD correlation coefficients from the 1000G European reference panel (53) and regional plots were generated by FUMA using LocusZoom (54) (**Supplementary Figure 2**). We verified that no variants were excluded due to the filtering on the 1000G reference panel; if a variant was filtered out and was within -/+ 250 kb of the top lead variant identified by FUMA, we considered it in this locus. However, if the variant was outside the -/+ 250 kb range, we considered it a new locus; these results are marked of * in the genomic locus tables (five in total). For the five loci we identified by manual search, we generated the LocusZoom plots for four (variant from the locus with the smallest p-value: rs1448813496, rs765377367, rs10993994 and rs201998020) using the reference panel generated from HUNT2-3 (**Supplementary Figure 2**). For rs191824624, we could not generate LocusZoom plot as the variant was not available in HUNT.

Overlapping loci between definitions were counted once and considered novel if not previously associated with UI.

We conducted the following downstream analyses using the top lead variants/loci identified by FUMA.

##### Gene-based, gene-set and tissue expression analyses

We performed MAGMA gene-based, gene-set and tissue expression analyses using FUMA. The genome-wide gene association analysis (gene-based analysis) was performed using MAGMA (v1.6) (55) and all variants located within protein-coding genes from Ensembl. The significance threshold was corrected for the number of protein-coding genes for which the input SNPs were mapped.

Further, we used MAGMA to perform gene-set analyses on the genes identified in the gene-based analysis, testing them with 17,023 genes set (Curated gene sets and Gene Ontology terms) from the MSigDB v7.0 database. The gene-set p-values were corrected for the 17,023 curated gene sets and GO terms tested (p-value < 2.94×10^-6^ after Bonferroni correction) To identify expression patterns of the prioritized genes across different tissues we performed MAGMA tissue specificity analysis using gene expression data from 53 specific tissue types in the Genotype Tissue Expression (GTEx) Project v8 (https://www.gtexportal.org/). The genes were prioritized using the results of the gene-based analysis. Tissues were identified as significant if their p-value < 9.43×10^-4^, Bonferroni correction for the 53 tissue types tested.

##### Cell-type analysis

We used cell-type specificity analysis as implemented in FUMA to identify cell types potentially involved in UI and its subtypes. Details about dataset selection are presented in the **Supplementary Text**.

##### Colocalization analysis

To test if UI loci may influence gene expression in relevant tissues, thereby suggesting a potential role of these genes in UI, we performed Bayesian colocalization using the R ‘coloc’ package (56) with summary statistics from the UI GWAS loci and publicly available eQTL data. We used Open Targets Genetics (https://genetics.opentargets.org/) to identify top lead variants with available eQTL data in relevant tissues within a gene mapped by FUMA, and selected these eQTLs and the corresponding GWAS loci for the analyses. Selection of the relevant tissues was done using the results of the MAGMA gene property analysis for tissue specificity and literature (**Supplementary Text**). We retrieved European ancestry eQTL data from the eQTL catalog (57) (GTEx v8, CommonMind, ROSMAP, FUSION and Alasoo 2018) using a publicly available API (function ‘request_association_from_api’) and adjusting parameters to obtain eQTL -/+ 500kb regions around the variant of interest (https://github.com/eQTL-Catalogue/eQTL-Catalogue-resources/blob/master/tutorials/API_v2/eQTL_API_tutorial.md). When eQTLs evidence for the same tissue but from two different datasets, we prioritized data from the Gtex v8. For some SNPs, eQTL evidence was identified in Open Targets Genetics but was not retrieved through the API. This may be due to the API accesses data from the eQTL Catalogue, whereas the eQTL evidence displayed in Open Targets Genetics may originate from the original datasets. The eQTL Catalogue reprocesses these datasets using standardized pipelines and additional quality control and filtering criteria, which may result in some associations not being included. In cases where the eQTL evidence was not available from eQTL Catalog but Open Targets showed evidence in a GTEx v8 dataset, we retrieved the corresponding data from GTEx v8 and performed colocalization. Overall, we tested 13 of the 14 unique GWAS loci for UI identified by Open Targets Genetics.

We conducted separate analyses for bladder tissue using GTEx v10; bladder tissue was not available in the eQTLCatalog (https://www.gtexportal.org/home/downloads/adult-gtex/qtl). We selected variants with eQTL evidence for the top lead variant of the GWAS in genes prioritized by FUMA (using significant variant-gene associations based on GTEx permutations). We identified one locus (chromosome 17) and considered the locus region and eQTL ±500 kb around the variant of interest. We considered *KANSL1-AS1* (not prioritized by FUMA) which is an antisense transcript of *KANSL1* (prioritized by FUMA). Given that bladder tissue is most relevant to UI, we conducted a sensitivity analysis restricting cis-eQTLs to ±50 kb around the lead variants to focus on local regulatory effects.

We set prior probabilities at 10⁻⁴ for phenotype or gene expression alone, and 10⁻⁶ for both (58). We considered posterior probabilities (PP4) > 75% as strong evidence of a shared causal variant for UI and gene expression. We used case-control (‘cc’) when formatting the data from the GWAS in coloc and reported the proportion of cases accordingly (‘s’).

##### Heritability and genetic correlation

We estimated SNP heritability at the liability scale using LD score regression (https://github.com/bulik/ldsc) (48,49) and estimated sample and population prevalences (**Supplementary Text**). Additionally, we estimated SUI-ICD and UUI-ICD pairwise genetic correlation. We restricted these analyses to SNP from HapMap3 and used LD scores derived from the 1000G European reference panel (https://alkesgroup.broadinstitute.org/LDSCORE/README_new_data_location.txt).

Additionally, we estimated genetic correlations between UI ICD code-based phenotypes and 2871 traits publicly available from the Complex Traits Genetics Virtual Lab server (18) using LD score regression from European reference panels (48,49). We reported genetically correlated phenotypes (p-value < 1.74×10^-5^, Bonferroni correction for 2,871 phenotypes). The phenotypes in genetic correlations with UI at r_g_> 0.4 were plotted by relevant categories; for clarity, we limited the plot at the 50 largest associations.

##### Phenome-wide association studies

We tested pleiotropic associations of the top lead variants identified in the meta-analyses (70 unique variants for ICD code-based definitions and three for self-reported definitions) with 1523 outcomes: 1326 ‘phecodes’ (ICD codes groupings representing clinically concepts for research (59)), 30 biomarkers and 167 continuous traits. We used publicly available GWAS summary statistics of European ancestry individuals (https://pan.ukbb.broadinstitute.org/). Results were considered significant if p-value < 4.50×10^-7^, Bonferroni correction for the 1523 phenotypes and the 73 variants.

##### UI drugs and gene target

To prioritize genes identified, we used DRUGBANK (https://go.drugbank.com/) to verify whether drugs currently used for UI (or in clinical trials) targeted the genes identified in our meta-analyze. We searched using the ”Indications” option for ‘stress urinary incontinence’, ‘urge urinary incontinence’, ‘urinary incontinence’ and ‘mixed urinary incontinence’. We selected the categories reported in the **Supplementary Table 24** in the column Indication (search ID).

##### PRS

We performed PRS-CS auto (60) for ICD code-based phenotypes leaving out HUNT. We used the UK Biobank European LD reference panel and computed the score using Plink2 (https://github.com/lcstoshio/tutorial_PRSCS_PRSCSx/blob/main/tutorial_PRScs.md and https://github.com/globalbiobankmeta/PRS/blob/main/run_prscsx_pipe.md) (60). We used the P+T method to generate the PRS for phenotypes with minimum five loci identified to avoid non-normality. We ran a meta-analysis excluding HUNT and extracted effect size estimates for each top lead variant. PRSs in HUNT were the sum of the number of risk alleles (from dosage), weighted by its regression coefficient (β) as implemented in Plink2 linear scoring. For both methods, we standardized the score to a normal distribution and estimated the associations between the PRS and UI in HUNT using logistic regression. Further, we estimated the variance explained by the PRS using the difference of Nagelkerke *R*^2^ estimates between a full model (PRS and covariates [age, sex, PCs and genotyping batch]) and a null model (covariates). We plotted UI prevalence by PRS deciles using the PRS-CS results.

##### Mendelian Randomization

We conducted two-sample MR to investigate the causal effects of six previously reported risk factors for UI: smoking status (14), alcohol consumption (36), BMI (15,16), parity (14,17) and pelvic organ prolapse for females, and benign prostatic hyperplasia for males (15). SNP-exposure association estimates were obtained from GWAS summary statistics of European ancestry populations from publicly available data (17,61–64). For sex-specific analyses and when available, we used sex-specific SNP-exposure association estimates.

Genetic instruments were derived by clumping (r² < 0.001) the lead SNPs for each risk factor (reported in the tables of the included studies), except for BMI, where we used genome-wide summary statistics and selected independent SNPs (r² < 0.001) passing the genome-wide significance threshold (p < 5 × 10⁻⁸). SNP–outcome associations for the selected genetic instruments were extracted from the GWAS meta-analyses of AUI-ICD, SUI-ICD, UUI-ICD and MUI-ICD. After harmonizing the risk alleles, we performed IVW two-sample MR to estimate associations, reporting odds ratios with 95% confident intervals, F-statistics, R^2^, and Cochran’s Q for heterogeneity. We performed sensitivity analyses including simple median, weighted median, MR-Egger methods. For binary exposures, estimates were scaled to represent the risk increase in UI per doubling (2-fold increase) in the prevalence of exposure, by multiplying the beta estimates by 0.693 (65). Steiger filtering was applied to verify whether each SNP explained more variance in the exposure than in the outcome; when SNPs explained more variance in UI, we re-ran the IVW MR and sensitivity analyses excluding those instruments.

##### Statistics and reproducibility

Statistical analysis has been performed using R v.4.2.3-4.5.2 and FUMA v1.6.1-1.6.5. The definitions used are provided in **Supplementary Table 33** for ICD-based definitions and **Supplementary Text** for self-reported definitions. The descriptive characteristics of the individuals included per study are available **Supplementary Table 1**.

## Data Availability

All data produced in the present study are available upon reasonable request to the authors

## Ethics

Study participants in HUNT have given written consent for the use of their data for research. The Regional Ethics Committee (REC) for Medical and Health Research Ethics in Central Norway have approved the study (REC reference number: 2018/1622). The UK Biobank received written consent from all the participants. UK Biobank was approved by the National Health Service (NHS) Research Ethics Service (reference number 11/NW/0382). The analyses in MGI have been approved by the Institutional review Board of the University of Michigan Medical School (IRB IDs HUM00071298, HUM00148297, HUM00099197, HUM00097962, and HUM00106315). Study subjects in FinnGen provided informed consent for biobank research, based on the Finnish Biobank Act. Alternatively, separate research cohorts, collected prior the Finnish Biobank Act came into effect (in September 2013) and start of FinnGen (August 2017), were collected based on study-specific consents and later transferred to the Finnish biobanks after approval by Fimea (Finnish Medicines Agency), the National Supervisory Authority for Welfare and Health. Recruitment protocols followed the biobank protocols approved by Fimea. The Coordinating Ethics Committee of the Hospital District of Helsinki and Uusimaa (HUS) statement number for the FinnGen study is Nr HUS/990/2017. The FinnGen study is approved by Finnish Institute for Health and Welfare (permit numbers: THL/2031/6.02.00/2017, THL/1101/5.05.00/2017, THL/341/6.02.00/2018, THL/2222/6.02.00/2018, THL/283/6.02.00/2019, THL/1721/5.05.00/2019 and THL/1524/5.05.00/2020), Digital and population data service agency (permit numbers: VRK43431/2017-3, VRK/6909/2018-3, VRK/4415/2019-3), the Social Insurance Institution (permit numbers: KELA 58/522/2017, KELA 131/522/2018, KELA 70/522/2019, KELA 98/522/2019, KELA 134/522/2019, KELA 138/522/2019, KELA 2/522/2020, KELA 16/522/2020), Findata permit numbers THL/2364/14.02/2020, THL/4055/14.06.00/2020, THL/3433/14.06.00/2020, THL/4432/14.06/2020, THL/5189/14.06/2020, THL/5894/14.06.00/2020, THL/6619/14.06.00/2020, THL/209/14.06.00/2021, THL/688/14.06.00/2021, THL/1284/14.06.00/2021, THL/1965/14.06.00/2021, THL/5546/14.02.00/2020, THL/2658/14.06.00/2021, THL/4235/14.06.00/2021, Statistics Finland (permit numbers: TK-53-1041-17 and TK/143/07.03.00/2020 (earlier TK-53-90-20) TK/1735/07.03.00/2021, TK/3112/07.03.00/2021) and Finnish Registry for Kidney Diseases permission/extract from the meeting minutes on 4^th^ July 2019. The Biobank Access Decisions for FinnGen samples and data utilized in FinnGen Data Freeze 12 include: THL Biobank BB2017_55, BB2017_111, BB2018_19, BB_2018_34, BB_2018_67, BB2018_71, BB2019_7, BB2019_8, BB2019_26, BB2020_1, BB2021_65, Finnish Red Cross Blood Service Biobank 7.12.2017, Helsinki Biobank HUS/359/2017, HUS/248/2020, HUS/430/2021 §28, §29, HUS/150/2022 §12, §13, §14, §15, §16, §17, §18, §23, §58, §59, HUS/128/2023 §18, Auria Biobank AB17-5154 and amendment #1 (August 17 2020) and amendments BB_2021-0140, BB_2021-0156 (August 26 2021, Feb 2 2022), BB_2021-0169, BB_2021-0179, BB_2021-0161, AB20-5926 and amendment #1 (April 23 2020) and it’s modifications (Sep 22 2021), BB_2022-0262, BB_2022-0256, Biobank Borealis of Northern Finland_2017_1013, 2021_5010, 2021_5010 Amendment, 2021_5018, 2021_5018 Amendment, 2021_5015, 2021_5015 Amendment, 2021_5015 Amendment_2, 2021_5023, 2021_5023 Amendment, 2021_5023 Amendment_2, 2021_5017, 2021_5017 Amendment, 2022_6001, 2022_6001 Amendment, 2022_6006 Amendment, 2022_6006 Amendment, 2022_6006 Amendment_2, BB22-0067, 2022_0262, 2022_0262 Amendment, Biobank of Eastern Finland 1186/2018 and amendment 22§/2020, 53§/2021, 13§/2022, 14§/2022, 15§/2022, 27§/2022, 28§/2022, 29§/2022, 33§/2022, 35§/2022, 36§/2022, 37§/2022, 39§/2022, 7§/2023, 32§/2023, 33§/2023, 34§/2023, 35§/2023, 36§/2023, 37§/2023, 38§/2023, 39§/2023, 40§/2023, 41§/2023, Finnish Clinical Biobank Tampere MH0004 and amendments (21.02.2020 & 06.10.2020), BB2021-0140 8§/2021, 9§/2021, §9/2022, §10/2022, §12/2022, 13§/2022, §20/2022, §21/2022, §22/2022, §23/2022, 28§/2022, 29§/2022, 30§/2022, 31§/2022, 32§/2022, 38§/2022, 40§/2022, 42§/2022, 1§/2023, Central Finland Biobank 1-2017, BB_2021-0161, BB_2021-0169, BB_2021-0179, BB_2021-0170, BB_2022-0256, BB_2022-0262, BB22-0067, Decision allowing to continue data processing until 31^st^ Aug 2024 for projects: BB_2021-0179, BB22-0067,BB_2022-0262, BB_2021-0170, BB_2021-0164, BB_2021-0161, and BB_2021-0169, and Terveystalo Biobank STB 2018001 and amendment 25^th^ Aug 2020, Finnish Hematological Registry and Clinical Biobank decision 18^th^ June 2021, Arctic biobank P0844: ARC_2021_1001.

## Data availability

The data are available in the supplementary and upon request. The summary statistics for the meta-analyses are available at NTNU Open Research Data (https://dataverse.no/dataverse/ntnu DOI: *will be added*) and in the GWAS catalog (accession number: *will be added*). The data used from the UK Biobank can be obtained by application (https://www.ukbiobank.ac.uk). The summary level data should be requested directly to FinnGen and MGI (for MGI, by emailing). The summary level data from NHS are available at https://www.ebi.ac.uk/gwas/efotraits/HP_0000020 (study accession numbers: GCST009926, GCST009927, GCST009928, GCST0010205). The eQTL data used are available at https://www.ebi.ac.uk/eqtl/ and https://www.gtexportal.org/home/downloads/adult-gtex/qtl (for the bladder only). The summary statistics used to perform the PheWas (biomarkers, continuous and phecode traits) are available at https://pan-dev.ukbb.broadinstitute.org/, we used the manifest and corresponding data from the 11 April 2022. The summary statistics used for the genetic correlation section are available in the Complex Traits Genetics Virtual Lab server (https://vl.genoma.io/).

## Code availability

The following publicly available tools have been used in this project:

R v.4.2.3-4.5.2, FUMA v1.6.1-1.6.5 (https://fuma.ctglab.nl/), SAIGE version 1.03 for GWAS using HUNT data and version 0.35.8.3 for GWAS using UK Biobank data (https://saigegit.github.io/SAIGE-doc/), METAL (https://genome.sph.umich.edu/wiki/METAL_Documentation), UCSC LiftOver command line tool (https://genome.ucsc.edu/cgi-bin/hgLiftOver), plink v2 (https://www.cog-genomics.org/plink/2.0/), Coloc (https://github.com/chr1swallace/coloc) as implemented following the tutorial eQTL Catalog resource tutorial (https://github.com/eQTL-Catalogue/eQTL-Catalogue-resources/blob/master/tutorials/API_v2/eQTL_API_tutorial.md), LDSC Score Regression (https://github.com/bulik/ldsc). The code we used will be made available on GitHub: *link*.

## Acknowledgement

We want to thank the participants and staff of The Trøndelag Health Study (HUNT), the UK Biobank, FinnGen and MGI.

The HUNT study is a collaboration between HUNT Research Center (Faculty of Medicine and Health Sciences, Norwegian University of Science and Technology [NTNU]), Trøndelag County Council, Central Norway Regional Health Authority, and the Norwegian Institute of Public Health. The genotyping in HUNT was financed by the National Institutes of Health; University of Michigan; the Research Council of Norway; the Liaison Committee for Education, Research and Innovation in Central Norway; and the Joint Research Committee between St. Olavs Hospital and the Faculty of Medicine and Health Sciences, Norwegian University of Science and Technology (NTNU). Analyses were performed in digital laboratories at HUNT Cloud, Norwegian University of Science and Technology (NTNU) (Trondheim, Norway).

This research has been conducted using the UK Biobank Resource under Application Number 40135.

We want to acknowledge the participants and investigators of the FinnGen study. The FinnGen project is funded by two grants from Business Finland (HUS 4685/31/2016 and UH 4386/31/2016) and the following industry partners: AbbVie Inc., AstraZeneca UK Ltd, Biogen MA Inc., Bristol Myers Squibb Inc. (and Celgene Corporation & Celgene International II Sàrl), Genentech Inc., Merck Sharp & Dohme LCC, Pfizer Inc., GlaxoSmithKline Intellectual Property Development Ltd., Sanofi US Services Inc., Maze Therapeutics Inc., Johnson&Johnson Innovative Medicine Inc., Novartis AG, Boehringer Ingelheim International GmbH and Bayer AG. Following biobanks are acknowledged for delivering biobank samples to FinnGen: Auria Biobank (www.auria.fi/biopankki), THL Biobank (www.thl.fi/biobank), Helsinki Biobank (www.helsinginbiopankki.fi), Biobank Borealis of Northern Finland (https://www.ppshp.fi/Tutkimus-ja-opetus/Biopankki/Pages/Biobank-Borealis-briefly-in-English.aspx), Finnish Clinical Biobank Tampere (www.tays.fi/en-US/Research_and_development/Finnish_Clinical_Biobank_Tampere), Biobank of Eastern Finland (www.ita-suomenbiopankki.fi/en), Central Finland Biobank (www.ksshp.fi/fi-FI/Potilaalle/Biopankki), Finnish Red Cross Blood Service Biobank (www.veripalvelu.fi/verenluovutus/biopankkitoiminta), Terveystalo Biobank (www.terveystalo.com/fi/Yritystietoa/Terveystalo-Biopankki/Biopankki/) and Arctic Biobank (https://www.oulu.fi/en/university/faculties-and-units/faculty-medicine/northern-finland-birth-cohorts-and-arctic-biobank). All Finnish Biobanks are members of BBMRI.fi infrastructure (https://www.bbmri-eric.eu/national-nodes/finland/). Finnish Biobank Cooperative -FINBB (https://finbb.fi/) is the coordinator of BBMRI-ERIC operations in Finland. The Finnish biobank data can be accessed through the Fingenious^®^ services (https://site.fingenious.fi/en/) managed by FINBB.

The authors acknowledge the Michigan Genomics Initiative participants, AI & Digital Health Innovation at the University of Michigan, and the University of Michigan Medical School Data Office for Clinical and Translational Research for providing data storage, management, processing, and distribution services. The authors acknowledge the Michigan Genomics Initiative participants, AI & Digital Health Innovation at the University of Michigan, the University of Michigan Medical School Central Biorepository, and the University of Michigan Advanced Genomics Core for providing data and specimen storage, management, processing, and distribution services, and the Center for Statistical Genetics in the Department of Biostatistics at the School of Public Health for genotype data curation, imputation, and management in support of the research reported in this publication.

BMB is supported by grants from the Liaison Committee for Education, Research and Innovation in Central Norway and the Joint Research Committee between St. Olavs Hospital and the Faculty of Medicine and Health Sciences, Norwegian University of Science and Technology (NTNU).

BNW is supported from the European Union’s Horizon Europe research and innovation programme under the Marie Skłodowska-Curie grant agreement No 101110878 and from the European Union’s Horizon 2020 research and innovation programme under grant agreement No 101016775. N.M.

WZ was supported by the National Human Genome Research Institute of the National Institutes of Health under Award Number R01HG014518. The content is solely the responsibility of the authors and does not necessarily represent the official views of the National Institutes of Health.

